# Loss of Function of the Cytoplasmic Fe-S Assembly Protein CIAO1 Causes a Neuromuscular Disorder with Compromise of Nucleocytoplasmic Fe-S Enzymes

**DOI:** 10.1101/2023.12.20.23300170

**Authors:** Nunziata Maio, Rotem Orbach, Irina Zaharieva, Ana Töpf, Sandra Donkervoort, Pinki Munot, Juliane Mueller, Tracey Willis, Sumit Verma, Stojan Peric, Deepa Krishnakumar, Sniya Sudhakar, A. Reghan Foley, Sarah Silverstein, Ganka Douglas, Lynn Pais, Stephanie DiTroia, Christopher Grunseich, Ying Hu, Caroline Sewry, Anna Sarkozy, Volker Straub, Francesco Muntoni, Tracey Rouault, Carsten G. Bönnemann

**Author notes:** These authors contributed equally to this work (Nunziata Maio, Rotem Orbach, Irina Zaharieva, Tracey Rouault, Carsten G. Bönnemann). Correspondence to: Carsten Bönnemann, MD, Full address: Neuromuscular and Neurogenetic Disorders of Childhood Section Neurogenetics Branch, National Institute of Neurological Disorders and Stroke NIH, Building 10, Room 2B39, 10 Center Drive, Bethesda, MD 20892-1477, USA. Correspondence to: Tracey Ann Rouault, MD.

## Abstract

Cytoplasmic and nuclear iron-sulfur enzymes that are essential for genome maintenance and replication depend on the cytoplasmic iron-sulfur assembly (CIA) machinery for cluster acquisition. Here we report that patients with biallelic loss of function in *CIAO1*, a key CIA component, develop proximal and axial muscle weakness, fluctuating creatine kinase elevation and respiratory insufficiency. In addition, they present with CNS symptoms including learning difficulties and neurobehavioral comorbidities, along with iron deposition in deep brain nuclei, macrocytic anemia and gastrointestinal symptoms. Mutational analysis and functional assays revealed reduced stability of the variants compared to wild-type CIAO1. Loss of CIAO1 impaired DNA helicases, polymerases and repair enzymes which rely on the CIA complex to acquire their Fe-S cofactors, with lentiviral restoration reversing all patient-derived cellular abnormalities. Our study identifies *CIAO1* as a novel human disease gene and provides insights into the broader implications of the iron-sulfur assembly pathway in human health and disease.

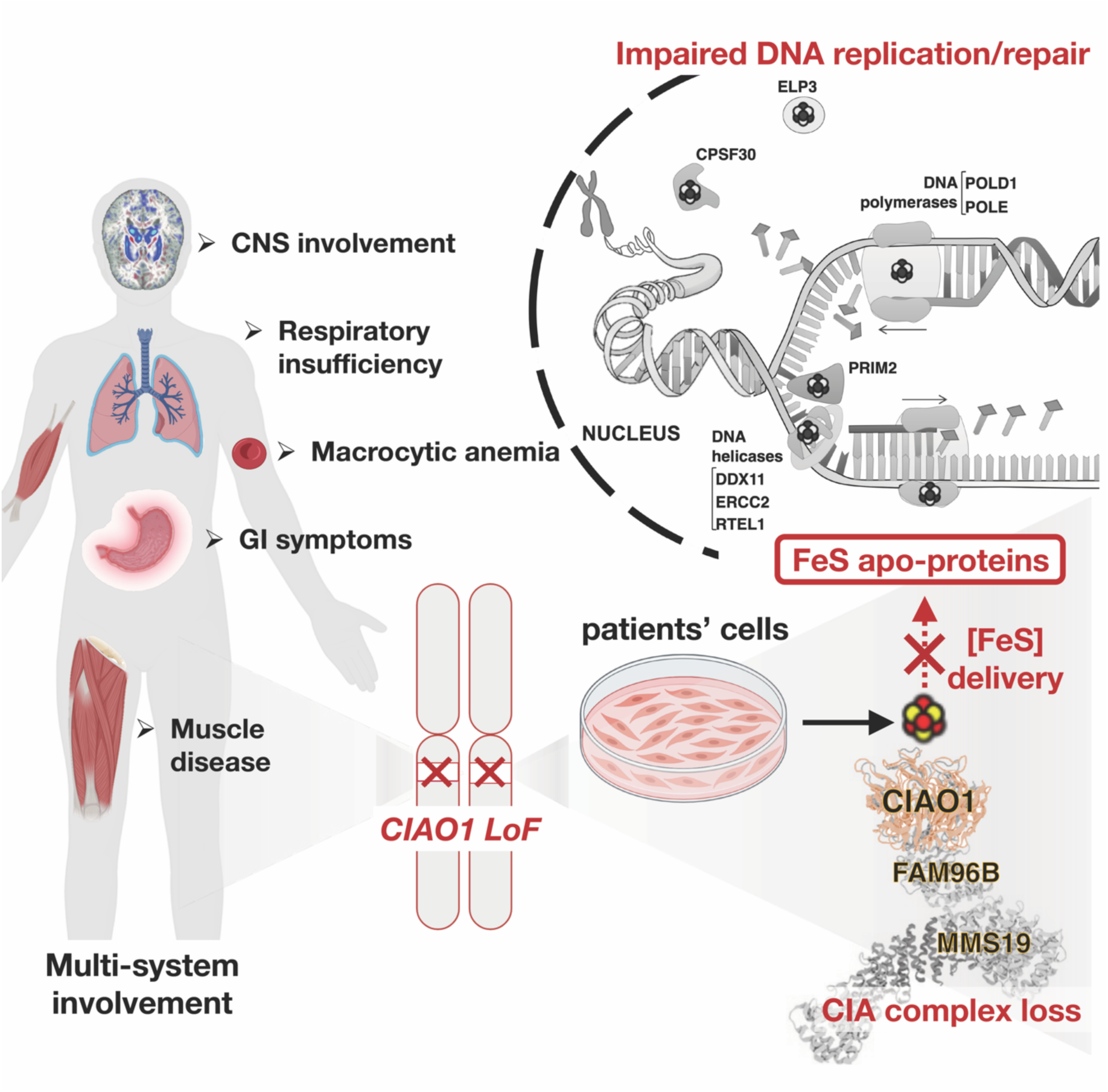
Graphical Abstract.

## Introduction

Iron-sulfur (Fe-S) clusters are ancient and evolutionarily conserved prosthetic groups with unique chemical properties that enable the proteins that contain them (Fe-S proteins) to function in several essential cellular pathways, including DNA replication and repair, ribosome biogenesis and protein translation, biosynthesis of heme and essential cofactors such as lipoic acid and biotin, electron transfer in the mitochondrial respiratory chain, and substrate coordination in dehydratases such as mitochondrial aconitase of the citric acid cycle.^1, 2^ Biogenesis of all Fe-S proteins depends on the core Fe-S cluster (ISC) assembly machinery (Fig. 1A). ^3, 4^ In mammalian cells, Fe-S clusters are assembled *de novo* by a complex composed of a cysteine desulfurase, NFS1, its binding partner, LYRM4 (also known as ISD11), the acyl carrier protein, NDUFAB1, the initial assembly scaffold, ISCU, an allosteric effector, frataxin, and ferredoxin, a source of reducing equivalents.^5, 6^ Upon assembly of a nascent cluster, the scaffold protein ISCU binds to an evolutionarily highly conserved co-chaperone/chaperone system, consisting of HSC20 (or HSCB) and HSPA9, respectively, in humans (Fig. 1A).^7, 8^ The HSC20-HSPA9 complex enhances Fe-S cluster transfer from the main scaffold ISCU directly to recipient proteins or to secondary carriers which then deliver Fe-S cofactors to recipient apoproteins. Importantly, the core mammalian ISC components have been previously identified in both the mitochondrial matrix and the cytosol and nucleus, where they operate in parallel to assemble Fe-S clusters in the subcellular compartments of multicellular eukaryotes.^7,9–14^

**Figure 1.**
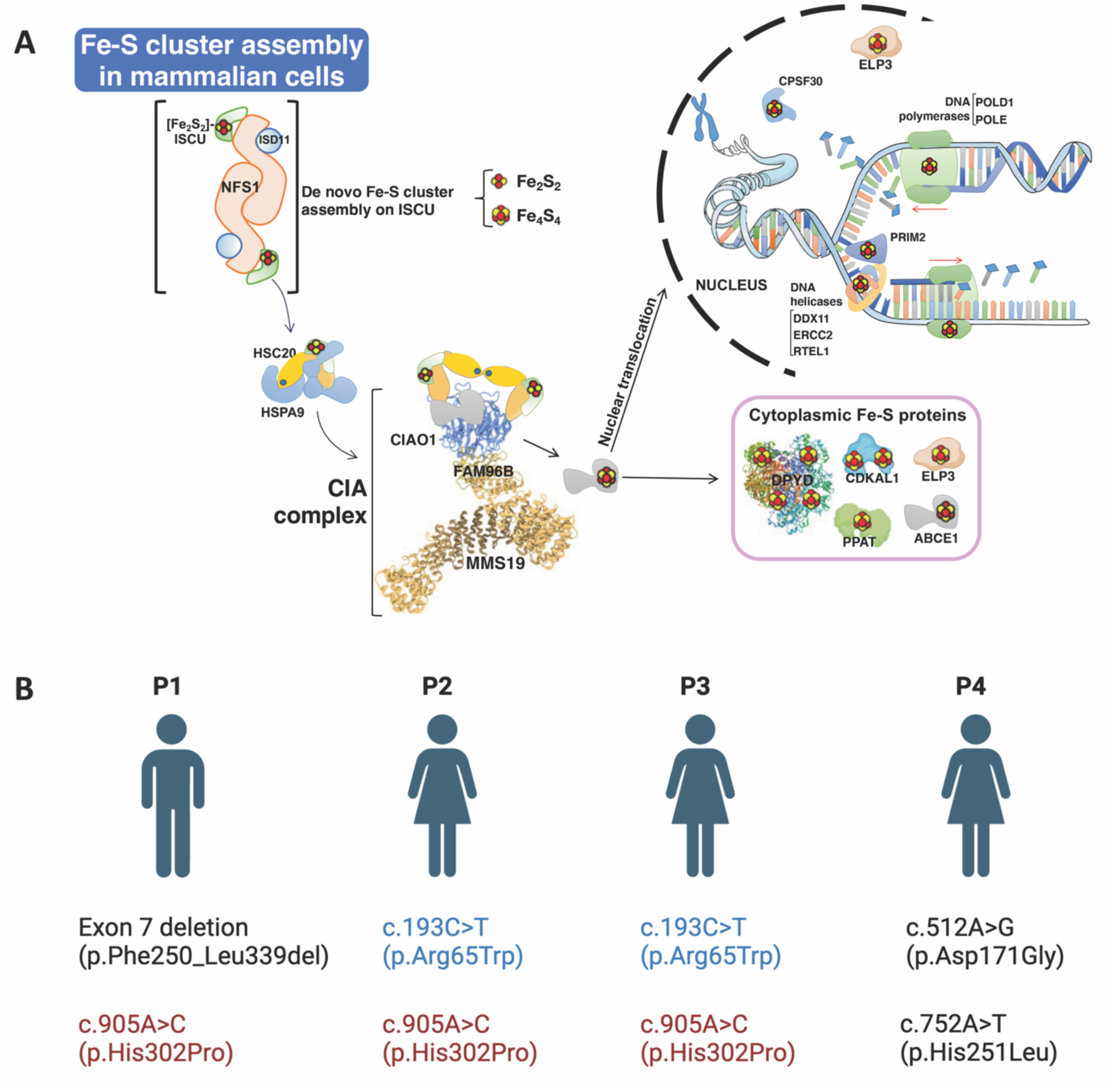
Identification of biallelic *CIAO1* variants in four independent patients with a neuromuscular condition of undefined etiology. A. Proposed model for the biogenesis of Fe-S clusters in the cytosol of mammalian cells. De novo assembly of Fe-S clusters occurs in the cytosol of mammalian cells upon the main scaffold protein ISCU by the coordinated action of a multi-protein complex, which consists of the cytosolic cysteine desulfurase NFS1 and the accessory protein ISD11. The HSC20/HSPA9 cochaperone/chaperone complex interacts with ISCU to facilitate cluster transfer to recipient proteins. The functional unit of HSC20 is a dimer^17^. A subset of recipient Fe-S proteins acquire their clusters directly from the HSC20/HSPA9/ISCU1 complex ^17^. Binding of HSC20 to the LYR motif of CIAO1 recruits the CIA targeting complex, which is known to form a platform to which Fe-S recipients involved in DNA metabolism dock to acquire their clusters ^15, 16^. The Fe-S proteins shown in the model were all identified as HSC20 interacting partners ^17^ (i.e. NUBP2, GLRX3, CIAPIN1, ABCE1, ERCC2, POLD1, PRIM2, PPAT, ELP3, CPSF30, DDX11, etc.). **B.** Diagram showing the biallelic *CIAO1* variants identified in patients. Recurring variants are shown in red and blue.

A subset of cytoplasmic and nuclear Fe-S enzymes requires the specialized cytoplasmic Fe-S assembly (CIA) complex, composed of CIAO1, MMS19 and FAM96B, which acts downstream of the ISC system, to acquire their cofactors (Fig. 1A).^15–17^ CIAO1 is a key component of the CIA machinery.^15–17^ The in-vivo functional consequences of loss of function mutations in *CIAO1* or in any of the CIA components have thus far remained unknown.

In this study, we identified four independent patients with biallelic loss of function in *CIAO1* who presented with a distinctive syndrome of predominantly neuromuscular manifestations and a spectrum of multisystemic findings. Biochemical and functional studies in patient-derived cell lines and tissues confirmed the pathogenicity of the variants and enabled the molecular characterization of the phenotype associated with loss of function in *CIAO1* characterized by the compromise of nucleocytoplasmic Fe-S enzymes involved in genome replication and maintenance, tRNA modification, and purine and pyrimidine metabolism. Lentiviral-mediated restoration of *CIAO1* expression reverted all the abnormalities of the patient-derived cells, thereby demonstrating that loss of function of CIAO1 caused the phenotype observed in the patients. Together, our studies define the critical requirement of *CIAO1* for human health and physiology and collectively contribute to a better understanding of the consequences of loss of function in a key component of the cytoplasmic Fe-S assembly pathway necessary for genome replication and maintenance.

## STAR Methods

### Recruitment and Samples Collection

We studied four unrelated patients with biallelic *CIAO1* variants, herein referred to as P1, P2, P3 and P4. All patients were followed in specialized neuromuscular clinics due to muscle weakness of unknown etiology. P1 was referred to the National Institutes of Health (NIH) by his neurologist. P2 was identified through the Matchmaker Exchange platform^18^, P3 and P4 were identified through the MYO-seq program^19^. Medical history and clinical evaluations, including muscle and brain magnetic resonance imaging (MRI) and muscle biopsies, were performed as part of the diagnostic efforts as standard diagnostic procedures. Laboratory testing, muscle biopsy histology slides and electron microscopy (EM) images/reports were independently reviewed. Patients’ variants in *CIAO1* were identified by whole-exome sequencing performed on whole-blood DNA obtained according to standard procedures. Samples for research-based testing, including blood (all patients), skin fibroblasts (P1, parents of P1, and P2) and muscle tissue (P1) were obtained via standard procedures, the muscle biopsy tissue was mounted in the gum guar oriented vertically, frozen in pre-cooled isopentane (2-methyl butane) and stored at –80 C before testing. Written informed consent and age-appropriate assent for research studies and procedures were obtained from the patients. Ethical approval was obtained for P1 via protocol 12-N-0095 approved by the National Institutes of Health Institutional Review Board, for P2 via the Great Ormond Street Hospital Research Ethics Committees GOSH 00/5802, and for both P3 and P4 via the National Research Ethics Service (NRES) Committee North East – Newcastle & North Tyneside 1 (reference 19/NE/0028).

### Exome, genome, and RNA Sequencing

P1 whole exome sequencing and analysis performed using Agilent Clinical Research Exome kit and Illumina HisSeq 2000 sequencing system with 100 bp paired-end reads and analyzed for sequence variants using a custom-developed analysis tool (Xome Analyzer, GeneDx). For P2, P3, and P4 whole exome sequencing and data processing were performed by the Genomics Platform at the Broad Institute of MIT and Harvard with a TWIST exome kit (P2) or with an Illumina Nextera (P3 and P4) and sequenced (150 bp paired reads) to cover >80% of targets at 20x and a mean target coverage of >100x. Exome sequencing data was processed through a pipeline based on Picard and mapping done using the BWA aligner to the human genome build 38. Variants were called using Genome Analysis Toolkit (GATK) HaplotypeCaller package version 3.5.

P1 human whole transcriptome sequencing on fibroblasts was performed by the Genomics Platform at the Broad Institute of MIT and Harvard. The transcriptome product combines poly(A)-selection of mRNA transcripts with a strand-specific cDNA library preparation, with a mean insert size of 550 bp. Libraries were sequenced on the HiSeq 2500 platform to a minimum depth of 50-75 million STAR-aligned reads. ERCC RNA controls are included for all samples, allowing additional control of variability between samples.

### Sashimi plots

Bam files were generated using the GTEXv10 pipeline (https://github.com/broadinstitute/gtex-pipeline) and aligned using reference genome GRCh38 (Gencode v39). Sashimi plots were generated using ggSashimi and a minimum splice junction threshold of 10 reads set.^20^ Control sample plots, when not separated, represent the mean junction reads of 3 aggregate samples.

### Cell culture methods

Dermal fibroblasts isolated from skin biopsies were grown in Dulbecco’s modified Eagle medium with 10% fetal bovine serum (Life Technologies) and 1% Penicillin/Streptomycin (Life Technologies) in 5% CO2 at 37°C.

### Lentiviral-mediated transduction of *CIAO1-V5* in patient-derived fibroblasts

Patient derived fibroblasts were engineered to stably express C-terminally V5-tagged *CIAO1* by lentiviral mediated transduction with pLENTI6.2/V5-DEST (Invitrogen). The ViraPower Lentiviral Expression System (Invitrogen) was used to produce viral particles harboring *CIAO1-V5* under the control of a CMV promoter, according to the manufacturer’s instructions. Briefly, pLENTI6.2/*CIAO1-V5* was cotransfected with the ViraPower Packaging Mix into HEK293T cells. The lentiviral stock collected 36 hours post-transfection was used to transduce P1– and P2– derived fibroblasts. Stable clones were established after six days of selection with blasticidin. Expression levels of *CIAO1*-V5 were assessed by western blot.

### Site-directed mutagenesis and transient transfection of plasmids encoding *CIAO1* variants in HeLa cells

Point mutations and deletion into *CIAO1* were introduced using the QuikChange II site-directed mutagenesis kit (Agilent Technologies), following the manufacturer’s instructions. All clones were verified for the insertion of the desired mutation by Sanger sequencing at Eurofins USA. Plasmid transfections into mammalian cells were performed with Lipofectamine 2000 (ThermoFisher Scientific), according to standard procedures.

### Subcellular fractionation into cytosol and mitochondria and immunoprecipitation experiments

Subcellular fractionation into cytosol and intact mitochondria was done as previously described ^7, 21, 22^. Briefly, mitochondria from patient-derived fibroblasts or HeLa cell pellets (∼10^9^ cells) were isolated from the cytosolic fractions after cell permeabilization with a buffer containing 0.1% digitonin in 210 mM mannitol, 20 mM sucrose, and 4 mM HEPES. The pellets after centrifugation at 700 x *g* for 5 min contained mitochondria, which were isolated by differential centrifugation and solubilized in lysis buffer I, containing 50 mM Bis-Tris, 50 mM NaCl, 10% w/v Glycerol, 0.001% Ponceau S, 1% lauryl maltoside, pH 7.2 and protease inhibitors.

The supernatants after the centrifugation at 700 x g containing soluble proteins were spun down at 21,000 x g for 20 min. The supernatant after the centrifugation was supplemented with a 1:1 volume (v/v) of a buffer containing 25 mM Tris, 200 mM NaCl, 1 mM EDTA, 1% NP-40 (pH 7.4) to obtain a final protein concentration of approximately 1 μg/μL. 500 mg of total cytosolic proteins were used for the Ips of CIAO1-V5 wild type and variants using agarose anti-V5 beads (Abcam # ab1229). Equilibrated anti-V5 agarose beads were added to the lysates and incubated for 2h at room temperature. Beads were extensively washed with lysis buffer, and bound proteins were eluted with Tris-Glycine pH 2.8 for 10 min at RT. The eluates were analyzed by SDS PAGE and immunoblot.

### Subcellular fractionation from muscle tissue sample

Snap-frozen muscle tissue specimens were ground to powder with mortar and pestle. Four volumes of extraction buffer (5 mM MOPS, pH 7.5 supplemented with 250 mM sucrose, 1 mM EDTA, and 0.1% ethanol) were added per gram of tissue, and samples were extracted by two sets of dounce strokes (20 strokes) in a pre-chilled dounce homogenizer, followed by incubation on ice for 10 min. The homogenate was centrifuged at 1000 x g for 10 min. The supernatant was transferred to a new tube and the pellet containing nuclei was washed with dilution buffer (5 mM MOPS, pH 8.0, supplemented with 1 mM EDTA and 0.1% ethanol), subsequently lysed in RIPA buffer and sonicated for the extraction of nuclear proteins. The supernatant after the first centrifugation step at 1000 x g was spun at 2000 x g for 10 min. The supernatant after the centrifugation 1000 x g was spun at 25,000 x g for 20 min and the pellet, containing a mixture of mitochondria, lysosomes, peroxisomes and ER membranes, was subjected to further purification by density gradient in Optiprep Density Gradient Medium (Sigma #D1556) diluted to the final required concentration in 5 mM MOPS, pH 8.0, containing 1 mM EDTA and 0.1% ethanol. The pellets containing light mitochondria were resuspended in 450 mL of 22.5% Optiprep Density Gradient Medium, layered between 200 mL of the 27.5% Optiprep solution (at the bottom) and 200 mL of the 20% Optiprep solution (on top). The gradient was centrifuged at 100,000xg for 1.5 hours. Mitochondria sedimented at the 22.5-27.5% interface and were lysed in 1.25x buffer I (50 mM BisTris, 50 mM NaCl, 10% w/v Glycerol, 0.001% Ponceau S, 1.2% Lauryl maltoside, pH 7.2, protease and phosphatase inhibitors). The supernatant after centrifugation at 21,000xg for 15 min was saved as mitochondrial lysates.

### DPYD activity assay

The dihydropyrimidine dehydrogenase (DPYD) activity was determined by thin layer chromatography (TLC), following a previously described protocol ^16, 23^, with the following modifications. Cytosolic cell lysates containing 150 μg of proteins isolated from patient-, parental– and control-derived fibroblasts or from muscle tissue lysates, as specified in the main text and figure legends, were applied to 50 μl of a reaction mix containing 25 mM Tris-HCl (pH 7.5), 0.1% digitonin, 2.5 mM MgCl_2_, 2mM DTT, 10 μM [4-^14^C]-thymine (0.1mCi/ml Moravek Inc. CA, USA), 10 mM NADPH. After 4 hours of incubation at 32 °C, the reaction was stopped by addition of 10 μl of perchloric acid (10% v/v). Reaction mixtures were centrifuged at 20,000 x g for 5 minutes and the supernatants analyzed by TLC.

### Native PAGE (BN-PAGE) analyses of mitochondrial respiratory complexes

The Native PAGE Novex Bis-Tris gel system (Thermo Fisher Scientific) was used to assess activities and levels of mitochondrial respiratory chain complexes, with the following modifications: only the Light Blue Cathode Buffer was used; 20 mg of membrane protein extracts were loaded/well; the electrophoresis was performed at 150 V for 1 h and 250 V for 2h.

### Complex I, complex II and complex IV in-gel activity assays and native immunoblots

In-gel Complex I, Complex II and Complex IV activity assays were performed as previously described ^7, 21, 22^. For Complex I activity, after resolution of the respiratory complexes by BN-PAGE, the gel was incubated with 0.1 M TrisCl, pH 7.4, containing 1 mg/ml nitrobluetetrazolium chloride (NBT) and 0.14 mM NADH at room temperature for 30-60 min. For Complex II, detection of succinate CoQ-reductase activity (SQR) (CoQ-mediated NBT reduction) was performed by incubating the gel for 30 minutes with 84 mM succinate, 2 mg/ml NBT, 4.5 mM EDTA, 10 mM KCN, 1 mM sodium azide and 10 mM ubiquinone in 50 mM PBS, pH 7.4. For complex IV, the gel was incubated in 50 mM phosphate buffer pH 7.4 containing 1mg/ml DAB (3,3’-diaminobenzidine) and 1mg/ml cytochrome c at room temperature for 30-45 min.

For the native immunoblots, PVDF was used as the blotting membrane. The transfer was performed at 25 V for 4 h at 4 °C. After transfer, the membrane was washed with 8% acetic acid for 20 min to fix the proteins, and then rinsed with water before air-drying. The dried membrane was washed 5-6 times with methanol (to remove residual Coomassie Blue G-250), rinsed with water and then blocked for 2h at room temperature in 5% milk, before incubating with the desired antibodies diluted in 2.5% milk overnight at 4 °C. In order to avoid strip and reprobe of the same membrane, which might allow detection of signals from the previous Ibs, samples were loaded and run in replicates on adjacent wells of the same gel and probed independently with different antibodies.

### Aconitase in-gel activity assay

Aconitase activity assay was performed as previously described ^24^.

### Inductively coupled plasma mass spectrometry (ICP-MS)

Iron content in the patient– and parental-derived fibroblasts was determined by ICP-MS (Agilent model 7900). 200 µL of concentrated trace-metal-grade nitric acid (Fisher) was added to isolated mitochondria and the organelles were digested overnight at 85 °C. Each sample was analyzed by ICP-MS after diluting with 3.8 mL deionized water.

### Immunoblots

Antibodies in this study were used at 1:1000 dilution unless otherwise specified and were as follows: anti-IRP1 antibody was prepared against purified human IRP1 and used at 1:5000 dilution. Anti-IRP2 antibody was prepared against a peptide covering the amino acid residues 137–209 of human IRP2 and used at 1:2000 dilution. Anti-ACO2 rabbit polyclonal antibody was raised against the synthetic peptide YDLLEKNINIVRKRLNR. Anti-TFRC antibody was from Thermo Scientific. Anti-FTH, FTL, NDUFS1, NDUFS8, NDUFV1, SDHA, SDHB, MTCO1, MTCO2, UQCRC1, UQCRC2, UQCRFS1, ATP5A, MT-CYB, MFN1/2 and total OXPHOS (Complex V, ATP5A subunit; Complex IV, COXII subunit; Complex III, UQCRC2 subunit; Complex II, SDHB; Complex I, NDUFB9 subunit) antibodies were from Abcam. Anti-CIAO1 and NFS1 were from Santa Cruz. Anti-tubulin, beta-actin, HSC20, HSPA9, FAM96A, ALAD were from Sigma. Anti-MMS19, FAM96B, ERCC2, ELP3, POLD1, PPAT, CDKAL1, CIAPIN1, GLRX3, DPYD, RTEL1, ABCE1, TOM20, FECH, MFN1 and CS were from Proteintech. Anti-BOLA2 was from Bethyl Laboratories. Anti-lipoate antibody was from EMD Millipore. Anti-OPA1 was from BD Biosciences.

### Radiolabeling experiments

The ^55^Fe incorporation assays were performed essentially as previously described ^25, 26^, with minor modifications. Patient– and parental-derived cell lines, as indicated, were grown in the presence of 1 μM ^55^Fe-transferrin (TF) for five to seven days. Transient transfection of C-terminally FLAG/MYC-tagged POLD1 for 16 hours was followed by subcellular fraction. Cytosolic extracts were subjected to immunoprecipitation with anti-FLAG agarose beads to immunocapture POLD1-FLAG. Samples collected after competitive elution (with 3X FLAG peptide at 100 mg/ml) were run on a native gel, followed by autoradiogram.

Alternatively, ^55^Fe incorporation into POLD1-FLAG/MYC was measured by scintillation counting of M2 FLAG beads (Sigma #A2220) after immunoabsorption of POLD1-FLAG/MYC, followed by extensive washes with buffer I. The background, corresponding to ^55^Fe measurements of eluates after anti-FLAG immunoprecipitations on cytosolic extracts from cells transfected with the empty vector was subtracted from each reading.

### Statistical Analyses

Where applicable, pairwise comparisons between two groups were analyzed using the two-tailed unpaired Student’s *t* test. Significance for multi-group comparisons was analyzed with two-way ANOVA followed by Sidak’s multiple comparisons test. All tests were performed with GraphPad Prism 9, and data were expressed as mean ± SD or SEM, as specified in the relative figure legends.

## Data availability

All data needed to evaluate the conclusions of the paper are present in the main text and Supplemental Information file. Therefore, all data are readily available to be shared with the appropriate data sharing agreements. There are no exceptions to the sharing of data, materials and softwares.

## Results

Loss of function in nearly all Fe-S assembly components has emerged over the past decade as the leading cause of multiple rare human conditions ^27^. However, thus far, no mutations have been found to induce deficiencies in the core components of the CIA complex composed of CIAO1, MMS19, and FAM96B. In this study, we present the case of four genetically unrelated individuals with loss of function in *CIAO1* (hereby named P1, P2, P3 and P4).

### Genetic findings

In all families, extensive next-generation-based sequencing did not identify pathogenic variants in the known human disease genes evaluated with adequate coverage. Upon further analysis of exome sequencing data, biallelic variants in *CIAO1* (NM_004804.3) were identified (Fig. 1B). Segregation testing when available (P1, P2) was consistent with autosomal recessive inheritance. Two missense variants were recurrent: p.His302Pro (*n* = 3, in P1, P2 and P3) and p.Arg65Trp (*n* = 2, in P2 and P3). Two variants were found in compound heterozygosity in P4 (p.Asp171Gly; p.His251Leu). P1 was compound heterozygous for a maternally inherited recurring missense variant (p.His302Pro) and a paternally inherited deletion of exon 7 (p.F250_L339del) of *CIAO1* (Fig. 1B), resulting in an out-of-frame transcript predicted to undergo nonsense mediated decay. This was confirmed by RNA sequencing on RNA extracted from dermal fibroblasts which revealed that P1 was apparently “homozygous” for the maternal *CIAO1* allele with no evidence of paternal reads (Fig. S1A, B). The Sashimi plot showed reduced *CIAO1* expression in P1’s fibroblasts compared to three aggregated control samples, with no effect on splicing (Fig. S1C).

The four *CIAO1* missense variants that were predicted to alter a conserved single amino acid were only found in heterozygosity in gnomAD (SVs v2.1).^28^ Variants were predicted to be disease causing by various in silico computational predictions (Table S1).

### Clinical presentation

All four patients had onset of weakness in early childhood to adolescent years (Fig. 2). Core phenotypic features in all patients consistent with the presence of a myopathy with dystrophic features included slowly progressive muscle weakness of proximal and axial skeletal muscles, mild facial and bulbar weakness, respiratory insufficiency (n= 4, FVC 51 to 63% predicted), fatigability/low endurance (n= 4), joint hyperlaxity (n= 4), ankle tightness (n= 4), calf pseudohypertrophy (n= 3, not commented for P2) and elevated CK (n= 4). Findings pointing to CNS and multisystemic involvement included learning disabilities/difficulties (n= 4), neurobehavioral comorbidities (n= 2), mild macrocytic anemia (n= 2), constipation and gastrointestinal symptoms (n= 2), obstructive sleep apnea (n= 2), and obesity (n= 3). There was no evidence of cardiac, neuropathic, ophthalmological, or hearing involvement.

**Figure 2.**
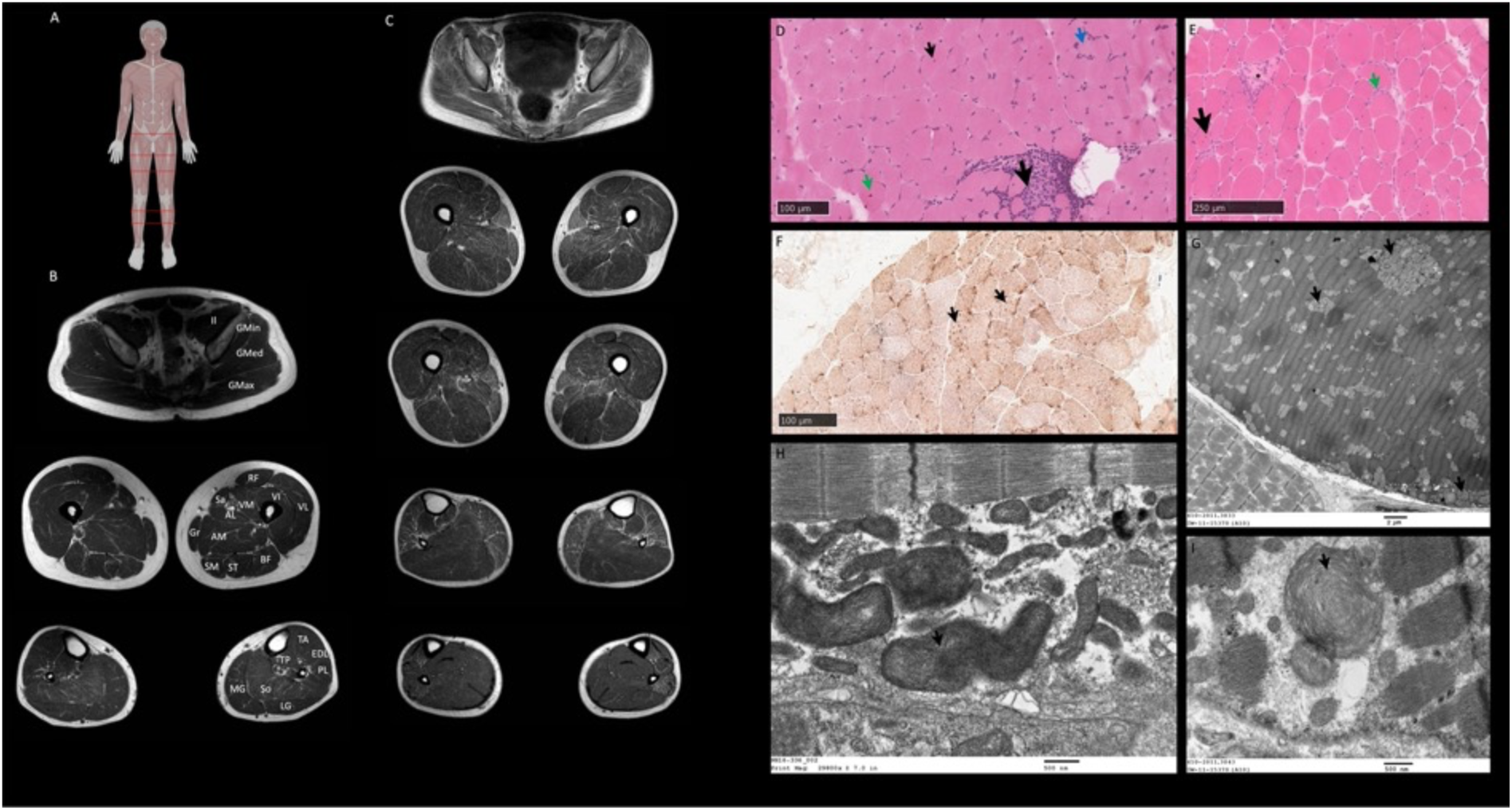
**Muscle MRI, muscle histopathology and muscle ultrastructural findings**. **A. Anatomical reference**. MRI scan positions are indicated with red lines on human reference image. **B. Normal muscle MRI**. T1 muscle MRI cross-sectional images showing normal anatomy of pelvic (top), thigh (middle) and lower leg (bottom) muscles. Abbreviations used to indicate specific muscle anatomy: AL, adductor longus; AM, adductor magnus; BF, biceps femoris; EDL, extensor digitorum longus; GMax, gluteus maximus; GMed, gluteus medius; Gmin, gluteus minimus; Gr, gracilis; Il, iliacus; LG, lateral gastrocnemius; MG, medial gastrocnemius; PL, peroneus longus; RF, rectus femoris; Sa, sartorius; SM, semimembranosus; ST, semitendinosus; VI, vastus intermedius; TP, tibialis posterior; VL, vastus lateralis; VM, vastus medialis. **C. Muscle MRI T1 sequence of P1.** Axial images (proximal to distal) at pelvic, thigh and calf levels showing mild diffuse fatty infiltration, greater in proximal muscles and slightly more pronounced in the posterior thigh and affecting the sartorius muscle more significantly. In the lower legs there is apparent hypertrophy of the soleus and gastrocnemius muscles and selective fatty infiltration of tibialis anterior, peroneus longus, peroneus brevis and medial gastrocnemius muscles. **D. Quadriceps muscle biopsy of P2. H&E** shows abnormal variation in fiber size, some internal nuclei (small arrow), occasional slight hypercontraction of fibers (green arrow), a focal area of cellularity possibly associated with necrosis (large arrow) and occasional slightly basophilic fibers (blue arrow). **E. Vastus lateralis muscle biopsy of P3. H&E** shows a wide abnormal variation in fiber size, occasional necrotic fiber (*), increased number of internal nuclei (black arrow), mild increase in endomysial connective tissue; and a little endomysial cellularity (green arrow). **F. Quadriceps muscle biopsy of P2, combined cytochrome oxidase (COX) and succinic dehydrogenase (SDH).** COX-SDH shows prominent mitochondria in several type 1 myofibers (arrows). **G. Electron microscopy (EM) of vastus medialis muscle biopsy of P1** shows scattered clusters of morphologically abnormal mitochondria (arrows). **H. EM of quadriceps muscle biopsy of P2** shows ultrastructural evidence of morphologically abnormal, large mitochondria with whorled cristae (arrow). **I. Additional EM from P1** demonstrates large mitochondria with disoriented cristae with concentric arrangements (arrow).

### Muscle diagnostics: imaging findings and biopsy analyses

#### Imaging findings

Muscle ultrasound (P1) and muscle MRI (P1, P2, P3, P4) uncovered a distinctive pattern of muscle involvement correlating with neuromuscular examination findings. Ultrasound revealed increased echogenicity of muscles in the upper and lower extremities (proximal > distal) and of paraspinal muscles. The lateral gastrocnemius and soleus muscles were hypertrophied, while the medial gastrocnemius was atrophied and markedly echogenic (Fig. 2A-C). Muscle MRI showed mild, diffuse fatty infiltration, greater in proximal muscles (all patients) and slightly more pronounced in the posterior thigh compartment (P1) (Fig. 2C) and affecting more significantly the sartorius (P1, P2, P3). In the lower extremities, there was hypertrophy of the soleus and gastrocnemius muscles, particularly the lateral gastrocnemius (P1, P2), and more selective fatty infiltration of specific muscles including the medial gastrocnemius (all patients), tibialis anterior, and peroneus muscles (P1, P2) (Fig. 2C).

#### Histopathological and ultrastructural findings

In all four patients, hematoxylin and eosin (H&E) stained sections demonstrated evidence for a myopathy with dystrophic features including variation in fiber size and an increase in internalized nuclei (Fig. 2D-E), as well as scattered degenerating/regenerating fibers and a mild to minimal increase in endomysial fibrosis (Fig. 2D-E). Foci of infiltrating immune cells, mainly macrophages (Fig. 2D, large arrow) were noted in all four cases. Upregulation of MHC-I was also noted, when assessed (P2, P3). Punctate material was present in a cytoplasmic distribution in mainly type I myofibers (Fig. 2F). The material strongly stained for cytochrome c oxidase (COX), succinic dehydrogenase (SDH) or combined COX-SDH (Fig. 2F) and thus corresponded to mitochondria.

Electron microscopy (EM) was performed on P1 and P2 skeletal muscle and revealed morphologically abnormal mitochondria, often in clusters, enlarged and elongated with aberrant cristae ultrastructure (Fig. 2G-I).

### Brain MRI findings

Brain MRI scans (P1, P2, P3) did not reveal any significant structural or white matter abnormalities. Notably, two patients (P1, P2) presented with increased iron deposition, beyond what would typically be expected for their age, based on findings of increased susceptibility on T2, susceptibility weighted imaging (SWI), and quantitative susceptibility mapping (QSM) sequences of the deep brain nuclei (globus pallidus, substantiate nigra, red nucleus, and dentate nucleus) (Fig. 3). The increased iron deposition has likely developed gradually, as it was not evident on earlier brain MRI scans for P1 and P2 (Fig. 3) ^29, 30^. Additional brain MRI findings were a single micro hemorrhage in the left middle frontal gyrus (P1), nonspecific right parietal white matter hyperintensities (P2), and mild diffuse cerebral and cerebellar volume reduction in brain MRI compared to an earlier scan (P2).

**Figure 3.**
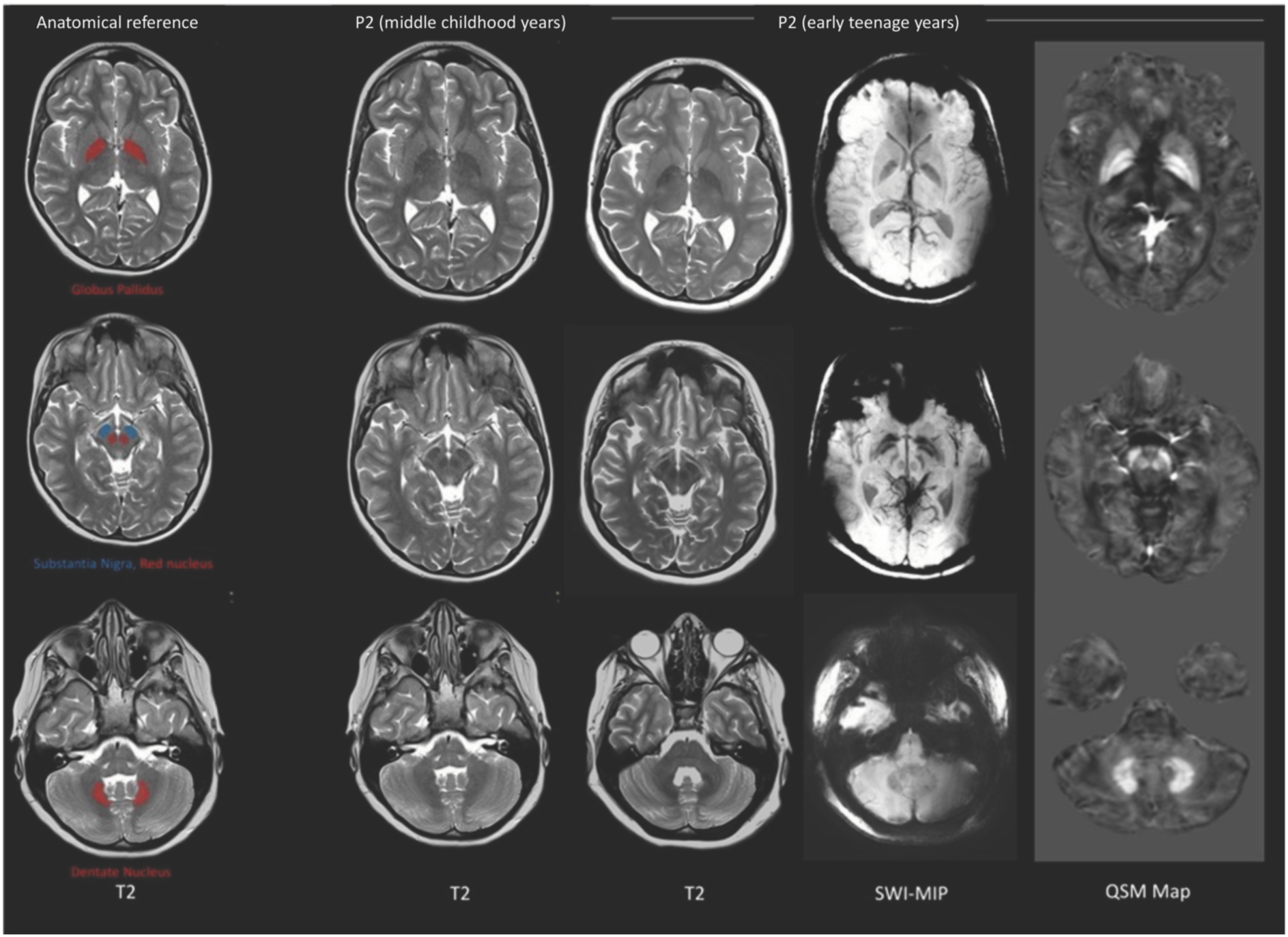
Brain MRI of P2 demonstrating evolving increased iron deposition in deep nuclei of the brain. Brain MRI performed in middle childhood years shows normal anatomy and susceptibility signals. Brain MRI acquired in early teenage years shows increased, atypical for age, susceptibility of bilateral globus pallidus (interna and externa with laminar sparing) [upper row], substantia nigra [middle row], red nucleus [middle row] and dentate nucleus [lower row]. The increased mineralization is demonstrated as hypointense signal on T2 and susceptibility weighted imaging (SWI), and hyperintensity on quantitative susceptibility mapping (QSM map). Of note, mild diffuse cerebral and cerebellar volume reduction is also apparent when compared to earlier scan. Left panel is displayed for anatomical reference.

### Biochemical and functional studies of CIAO1 variants identified in patients

#### The *CIAO1* variants identified in patients cause protein instability and compromised biogenesis of multiple Fe-S proteins that acquire their clusters from the CIA complex

The amino acid residues substituted in the patients are completely conserved across different species (Fig. S2A), and structural analysis revealed that they are located on the short loops that interconnect the β-propeller domains (also known as blades) of CIAO1 (Fig. S2B), likely playing critical roles in proper folding and stability of the protein. Particularly, the CIAO1 domain deleted in P1 (Phe250 through Leu339) is required to anchor CIAO1 to FAM96B in the overall architecture of the CIA complex (Fig. 4A, S2B). CIAO1 protein levels were profoundly diminished in cytosolic lysates from P1-derived fibroblasts compared to parental cells (Fig. 4B, S3A-B), with a concomitant loss of the known CIAO1-interacting partners MMS19 and FAM96B (Fig. 4B, S3A-B). Levels of the FAM96B paralogous protein, FAM96A, that has been shown to interact with CIAO1 but not with MMS19, ^31^ were also decreased in P1-derived lysates compared to parental cells (Fig. 4B). Following the loss of the three core components of the CIA complex, which specifically localizes to the cytosolic compartment of mammalian cells (Fig. S3A-B), levels of several Fe-S proteins that are known clients of the CIA machinery were decreased in lysates from P1 compared to parental cells (Fig. 4B-C). This observation is consistent with the reported instability of recipient Fe-S apoproteins when they fail to ligate their cofactors. ^21, 22, 32, 33^ Levels of the components of the *de novo* ISC assembly machinery, NFS1, HSC20 and HSPA9, which localizes to both the cytosolic and mitochondrial compartments of mammalian cells, ^7, 9–14^ were unchanged in lysates from P1 compared to parental cell lysates (Fig. S3C-D). Functional assays revealed selective compromise of the biogenesis of Fe-S enzymes that are known clients of the CIA complex, namely DPYD (Fig. 4D), which is involved in pyrimidine base degradation, and POLD1, responsible for replication of the DNA lag strand (Fig. 4E-F).

**Figure 4.**
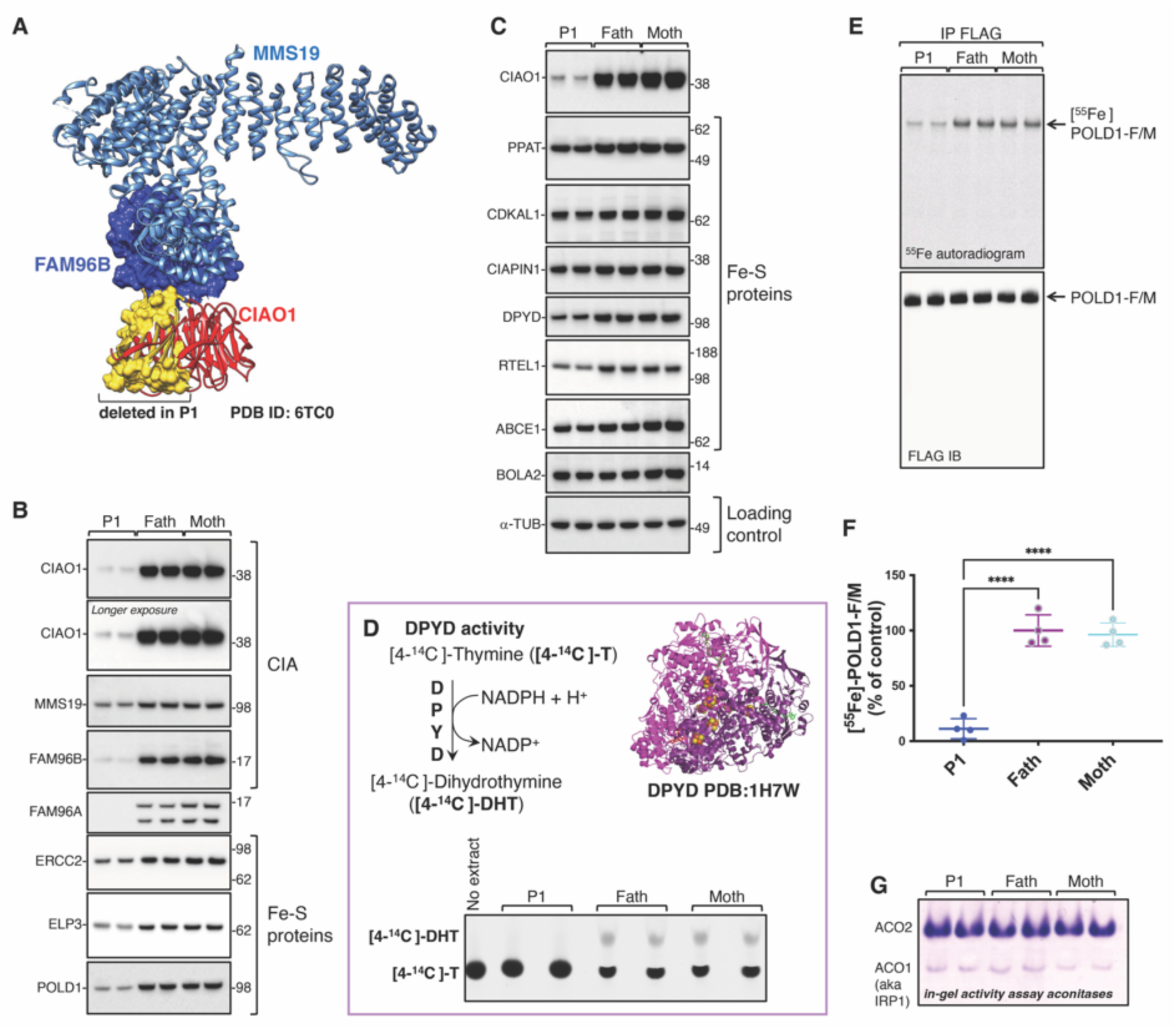
The *CIAO1* variants identified in P1 cause protein instability and compromised biogenesis of multiple Fe-S clients of the CIA complex. A. Schematic of the segregation inheritance of the CIAO1 variants identified in P1. **B.** 3D structure of the CIA complex consisting of CIAO1, MMS19 and FAM96B (PDB ID: 6TC0). CIAO1 is shown in ribbon– and surface-mode representations and colored in red, except for the domain with the amino acid residues that are deleted in P1, which is rendered in surface-mode representation and colored in yellow. An arrow indicates the location of His302 in CIAO1, a residue that is substituted by proline in P1. **C.** and **D.** Levels of the CIA components and Fe-S recipient proteins in P1– and parental-derived fibroblasts. Levels of FAM96A, a known interacting partner of CIAO1 but not of MMS19 or FAM96B, are also shown, along with the cytosolic iron and Fe-S cluster chaperone, BOLA2^45^. Alpha-tubulin (a-TUB) is included as a reference for loading control. **E.** Top left corner, schematic representation of the reaction catalyzed by the cytosolic Fe-S enzyme DPYD. DPYD converts [4-^14^C]-thymine ([4-^14^C]-T) to [4-^14^C]-dihydrothymine ([4-^14^C]-DHT). Top right corner, ribbon representation of the crystal structure of DPYD (PDB ID: 1H7W), which assembles into a dimer, containing a total of 8 [Fe_4_S_4_] clusters. Bottom section, DPYD-mediated conversion of [4-^14^C]-thymine to [4-^14^C]-dihydrothymine in lysates derived from P1 or control cells (the latter representing P1 parental fibroblasts), as indicated, assayed by thin layer chromatography (TLC) and autoradiography. The reaction mix containing the substrate of the reaction [4-^14^C]-T without cell extract was loaded as a negative control (no extract) to visualize the substrate (4-^14^C-thymine) by TLC. **F.** Representative ^55^Fe incorporation into POLD1-FLAG/MYC transiently expressed for 16 h in fibroblasts derived from P1 and in parental fibroblasts. Anti-FLAG immunoblot (IB) to verify that equal amounts of POLD1-F/M were immunoprecipitated is also shown. **G.** Quantification of radioactive iron incorporated into POLD1-F/M as assessed by scintillation counter. [^55^Fe]-POLD1-F/M levels in control cells (father of P1) were quantified and set to 100%. Values are expressed as % of control and are given as mean ± standard error of the mean (SEM). p< 0.0001. (C-G, *n*=4 biological replicates). **H.** In-gel activity assays of cytosolic (ACO1) and mitochondrial (ACO2) aconitases demonstrated unaltered activities of the [Fe_4_S_4_] enzymes in fibroblasts from P1 compared to parental fibroblasts (control cells).

To assess the impact of the mutations on protein stability and function, we generated and recombinantly expressed in HeLa cells the *CIAO1* variants identified in the patients. All the variant CIAO1 proteins had profoundly diminished stability when compared to wild type CIAO1 (Fig. S3E) and failed to interact with the CIA and the de novo ISC assembly components, as well as to recruit recipient Fe-S proteins (Fig. S3F). Collectively, these findings strongly support that the identified variants are indeed pathogenic.

Consistent with the notion that the CIA machinery is a specialized complex responsible for the biogenesis of a large subset of cytoplasmic and nuclear Fe-S proteins but not all cytosolic enzymes, activity of cytosolic aconitase, ACO1, (also known as IRP1) was unaltered in P1-derived lysates compared to parental cell lysates (Fig. 4G, S4A-H), and there was no evidence of mitochondrial iron overload in mitochondria isolated from P1 fibroblasts (Fig. S4G).

To unequivocally demonstrate that the phenotype reported in the patient cells was due to loss of function in *CIAO1*, we performed lentiviral-mediated transduction of C-terminally V5-tagged *CIAO1* in P1-derived fibroblasts to restore expression of the wild type protein. We found that CIAO1-V5 fully reverted all the abnormalities of the patient cells; specifically, it restored levels of the CIA components, MMS19 and FAM96B, and of FAM96A, and stabilized recipient Fe-S proteins that acquire their clusters from the CIA complex (Fig. 5A). Moreover, functional assays demonstrated normalized radioactive iron incorporation into POLD1 in patient-derived cells upon re-expression of CIAO1-V5 (Fig. 5B-C), along with full restoration of DPYD activity (Fig. 5D).

**Figure 5.**
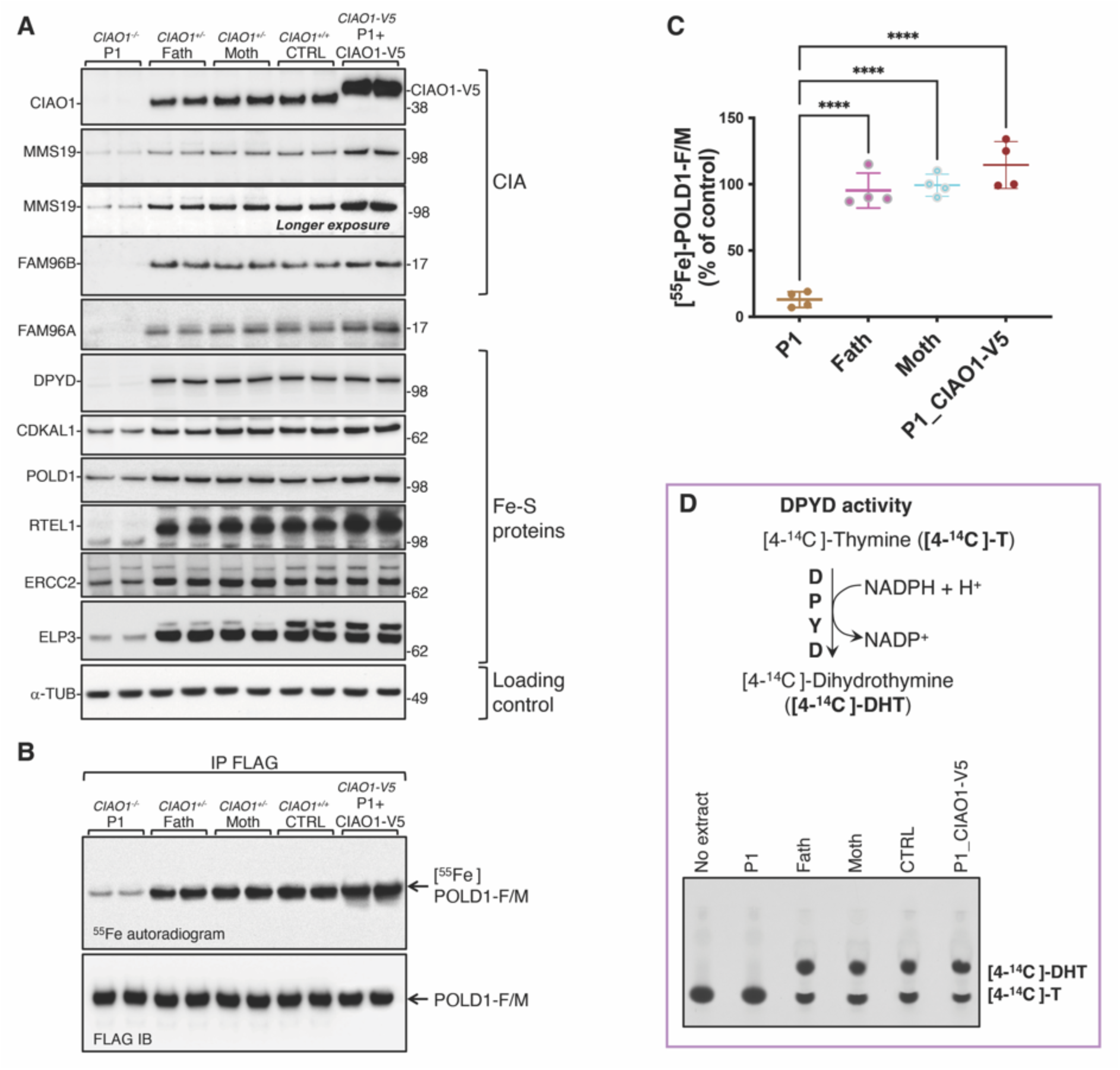
Lentiviral-mediated transduction of V5-tagged *CIAO1* wild type in patient-derived cells restored levels of the CIA components and stabilized recipient Fe-S proteins while restoring the activity of DPYD and radioactive iron incorporation into recombinantly expressed FLAG/MYC-tagged POLD1. A. Immunoblots to CIAO1, MMS19, FAM96B, FAM96A, and recipient Fe-S proteins (DPYD, CDKAL1, POLD1, RTEL1, ERCC2, and ELP3) on lysates from P1-(*CIAO1^-/-^*), parental-(*CIAO1^+/-^*), control (CTRL; *CIAO1^+/+^*)-derived fibroblasts and from P1-derived fibroblasts after lentiviral-mediated restoration of *CIAO1* wild type (*CIAO1-V5)*. **B.** Representative ^55^Fe incorporation into POLD1-FLAG/MYC transiently expressed in P1-, parental-, control (FC2)-derived fibroblasts and in P1-derived fibroblasts after lentiviral-mediated restoration of *CIAO1* wild type (*CIAO1-V5)*. Anti-FLAG immunoblot (IB) to verify that equal amounts of POLD1-F/M were immunoprecipitated is also shown. **C.** Quantification of radioactive iron incorporated into POLD1-F/M as assessed by scintillation counter. [^55^Fe]-POLD1-F/M levels in control cells (father of P1) were quantified and set to 100%. Values are expressed as % of control and are given as mean ± standard error of the mean (SEM). p< 0.0001. (C, *n*=4 biological replicates). **D.** Schematic representation of the reaction catalyzed by the cytosolic Fe-S enzyme DPYD. DPYD converts [4-^14^C]-thymine ([4-^14^C]-T) to [4-^14^C]-dihydrothymine ([4-^14^C]-DHT). Bottom section, DPYD-mediated conversion of [4-^14^C]-thymine to [4-^14^C]-dihydrothymine in lysates derived from P1-, parental– or control-fibroblasts (representing a fibroblast cell line harboring two wild type copies of *CIAO1*, CTRL), and in P1-derived fibroblasts stably expressing *CIAO1-V5*, as indicated, assayed by thin layer chromatography (TLC) and autoradiography. The reaction mix containing the substrate of the reaction [4-^14^C]-T without cell extract was loaded as a negative control (no extract) to visualize the substrate (4-^14^C-thymine) by TLC.

A second primary fibroblast cell line derived from P2 was generated and biochemically characterized. Despite harboring only missense variants in CIAO1 (p.H302P/p.R65W), P2-derived fibroblasts exhibited a similar loss of CIAO1 protein as observed in P1-derived cells (Fig. 6A), along with compromised stability of FAM96A and several cytoplasmic and nuclear Fe-S enzymes (Fig. 6A). Lentiviral-mediated restoration of *CIAO1* expression in P2-derived fibroblasts also restored levels of MMS19 and FAM96B (Fig. 6B-C), as well as stability of recipient Fe-S proteins (Fig. 6C). Both P1– and P2-derived fibroblasts did not exhibit a significant mitochondrial defect (Fig. 6D-E, S5A-K, S6A-C), indicating that mitochondrial function was not profoundly impaired upon loss of the CIA complex in fibroblasts (*vide infra* for muscle tissue results). Functional assays in P2-derived fibroblasts revealed compromised biogenesis of Fe-S enzymes that depend on the CIA complex for cluster acquisition. Specifically, radioactive iron incorporation into POLD1 was severely impaired to an extent comparable to that observed in P1 (Fig. 6F-G). Lentiviral-mediated re-expression of *CIAO1* wild type in P2 fibroblasts normalized ^55^Fe incorporation into POLD1 (Fig. 6F-G), confirming that loss of CIAO1 caused its compromised maturation.

**Figure 6.**
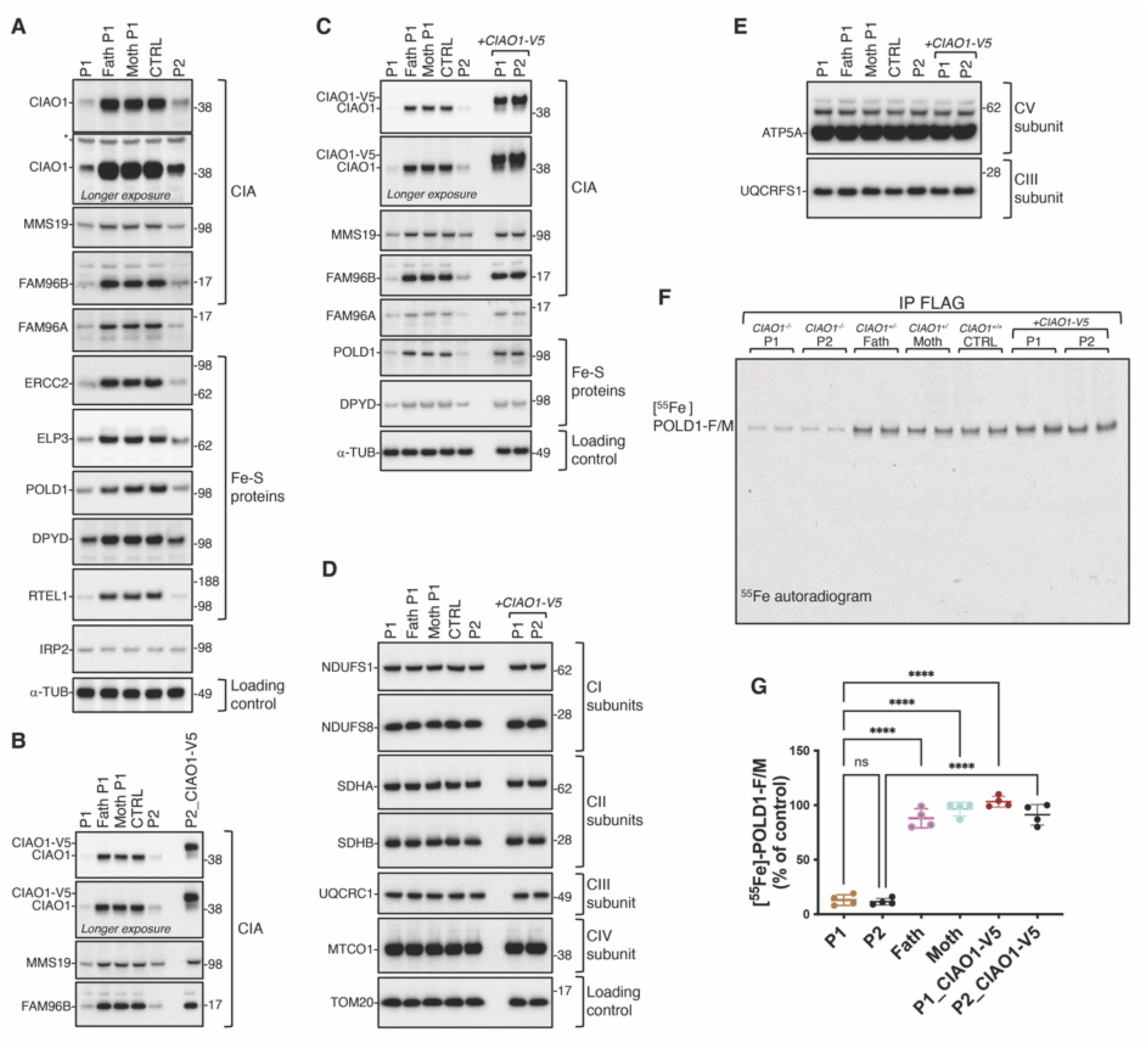
A second cell line derived from P2, who is not genetically related to P1 and harbors distinct missense mutations in *CIAO1*, demonstrates similar abnormal characteristics as observed in P1. These defects in P2-derived cells were entirely reversed when the wild-type *CIAO1* gene was reintroduced. A. SDS immunoblots to the CIA components, to FAM96A and to recipient Fe-S proteins (ERCC2, ELP3, POLD1, DPYD and RTEL1) in fibroblasts derived from P1 and his parents, from a control cell line harboring two wild type copies of *CIAO1*, and from P2, as indicated. Alpha tubulin (a-TUB) levels are shown as a reference for the loading control. **B.** SDS immunoblots to the CIA components in P1– and parental-derived fibroblasts, in a control cell line harboring two wild type copies of *CIAO1*, and in P2, as indicated. P2 cells lentivirally transduced with V5-tagged *CIAO1* wild type showed full restoration of the levels of CIA components. **C.** SDS immunoblots to CIA components, to FAM96A and to Fe-S proteins that are recipients of the CIA complex (POLD1, DPYD), as indicated, in P1– and parental-derived fibroblasts, in a control cell line harboring two wild type copies of *CIAO1*, and in P2, as indicated. Lysates from P1 and P2 cell lines lentivirally transduced with V5-tagged *CIAO1* wild type were also included and showed full restoration of the levels of CIA components and Fe-S recipients. Alpha tubulin (a-TUB) levels are also shown as a reference for the loading control. **D.** SDS immunoblots to subunits of the mitochondrial respiratory chain complexes I (NDUFS1, NDUFS8), II (SDHA, SDHB), III (UQCRC1) and IV (MTCO1) in lysates from P1– and parental-derived fibroblasts, from a control cell line harboring two wild type copies of *CIAO1*, and from P2, as indicated. Lysates from P1 and P2 cell lines lentivirally transduced with V5-tagged *CIAO1* wild type were also included. Levels of the mitochondrial marker TOM20 are shown as a reference for loading control. **E.** SDS immunoblots to the mitochondrial respiratory chain subunits of complex V (ATP5A) and complex III (UQCRFS1) in lysates from P1– and parental-derived fibroblasts, from a control cell line harboring two wild type copies of the *CIAO1* gene, and from P2, as indicated. Lysates from P1 and P2 cell lines lentivirally transduced with V5-tagged *CIAO1* wild type were also included. **F.** Representative ^55^Fe incorporation into POLD1-FLAG/MYC transiently expressed in P1-, parental-, control – derived fibroblasts and in P1– and P2-derived fibroblasts after lentiviral-mediated restoration of *CIAO1* wild type. **G.** Quantification of radioactive iron incorporated into POLD1-F/M as assessed by scintillation counter. [^55^Fe]-POLD1-F/M levels in control cells (father of P1) were quantified and set to 100%. Values are expressed as % of control and are given as mean ± standard error of the mean (SEM). p< 0.0001. (C, *n*=4 biological replicates).

### Patients’ skeletal muscle revealed compromise of the CIA complex activity and abnormal mitochondrial morphology and function

Because skeletal muscle is the most consistently affected tissue in our patients with biallelic *CIAO1* variants, we extended the biochemical analysis to muscle biopsy material, which was available from P1. We found that levels of CIAO1 and the other CIA components, MMS19 and FAM96B, were profoundly diminished in P1 skeletal muscle (Fig. 7A), with concomitant loss of several cytoplasmic and nuclear Fe-S proteins (Fig. 7A). Similar to what was observed in P1-derived fibroblasts, levels of iron regulatory proteins (IRP1 and IRP2) were unaltered in P1-derived muscle tissue lysates compared to control (Fig. 7B), whereas DPYD activity was severely impaired (Fig. 7C). The electron microscopy (EM) data collected for P1 and P2 were indicative of mitochondrial ultrastructural abnormalities in the patients’ skeletal muscle (Fig. 2G-I). Consistently, we observed compromised assembly and activities of the mitochondrial respiratory chain complexes. Specifically, levels of subunits of complexes I, II, III and IV were all decreased in P1 muscle mitochondria compared to control lysates (Fig. 7D), and activities and levels of fully assembled complexes I, II and IV were also decreased (Fig. 7E, 7F, respectively), pointing to a muscle specific mitochondrial dysfunction secondary to *CIAO1* loss of function.

**Figure 7.**
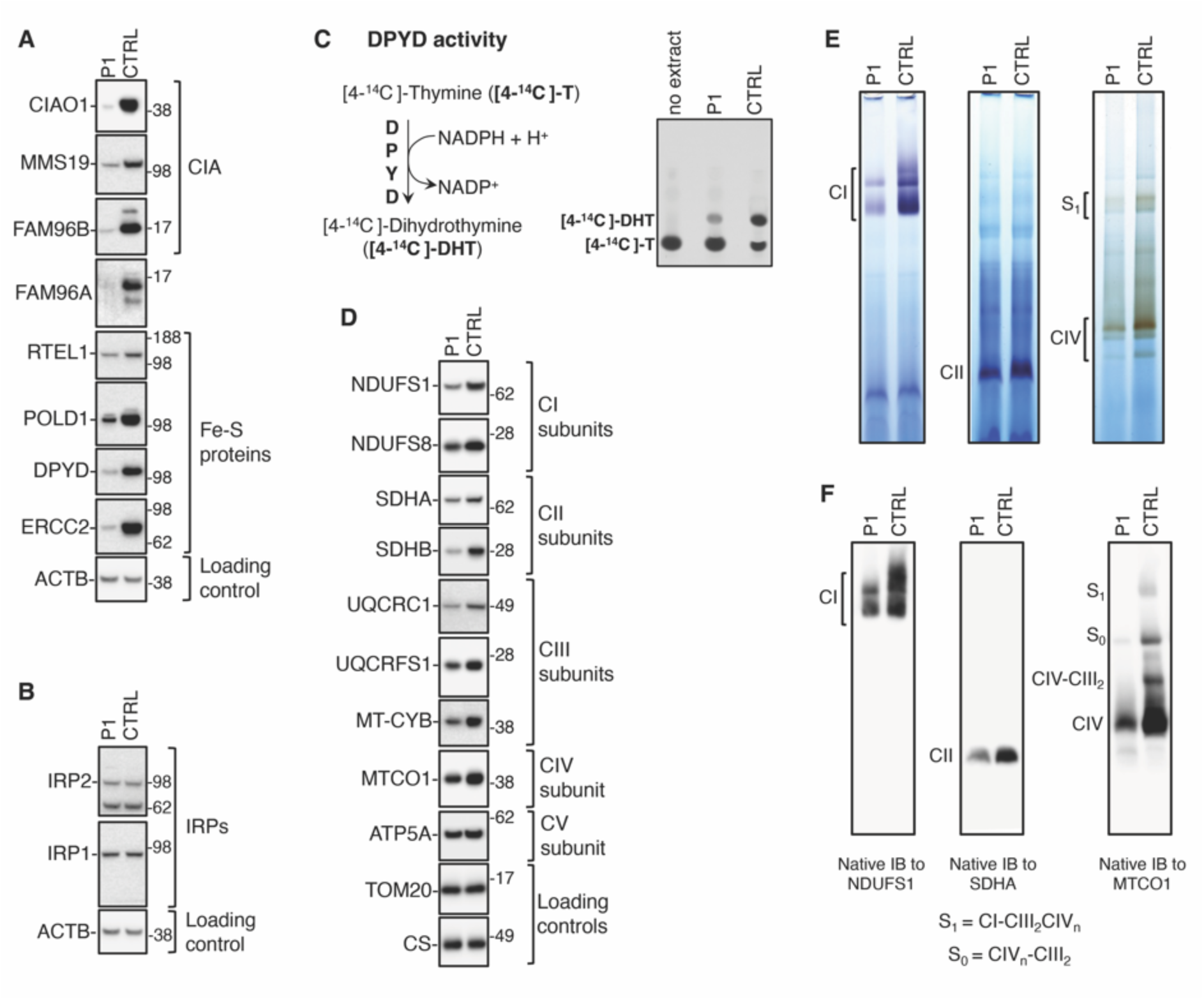
Mitochondrial dysfunction and compromised biogenesis of Fe-S recipients of the CIA targeting complex in muscle tissue from P1. A. SDS immunoblots to the CIA components (CIAO1, MMS19 and FAM96B) and to FAM96A in P1 and control (*CIAO1^+/+^*) muscle tissue specimens. Levels of cytoplasmic and nuclear Fe-S recipient proteins (RTEL1, POLD1, DPYD and ERCC2) were also assessed. b-actin (ACTB) was included as a reference for the loading control. **B.** SDS immunoblots to IRP1 and IRP2 in P1 and control (*CIAO1^+/+^*) muscle tissue specimens. **C.** On the left is a schematic representation of the reaction catalyzed by the cytosolic Fe-S enzyme DPYD. DPYD converts [4-^14^C]-thymine ([4-^14^C]-T) to [4-^14^C]-dihydrothymine ([4-^14^C]-DHT). On the right is the DPYD-mediated conversion of [4-^14^C]-thymine to [4-^14^C]-dihydrothymine in lysates derived from P1 and control (*CIAO1^+/+^*; CTRL) derived muscle tissue specimens, assayed by thin layer chromatography (TLC) and autoradiography. The reaction mix containing the substrate of the reaction [4-^14^C]-T without cell extract was loaded as a negative control (no extract) to visualize the substrate (4-^14^C-thymine) by TLC. **D.** SDS-IBs to subunits of mitochondrial respiratory complex I (NDUFS1 and NDUFS8), complex II (SDHA, SDHB), complex III (UQCRC1, UQCRFS1, MT-CYB), complex IV (MTOC1) and complex V (ATP5A) in P1– and control-derived muscle tissue specimens. Levels of TOM20 and citrate synthase (CS) were included as a reference for the loading control. **E.** In-gel activity assays of mitochondrial respiratory complexes I (CI), II (CII) and IV (CIV) in P1– and control-derived muscle tissue specimens. **F.** Native immunoblots (IBs) to subunits of complex I (NDUFS1), complex II (SDHA) and complex IV (MTCO1) to assess the overall levels of fully assembled respiratory complexes.

## Discussion

The in vivo pathophysiological consequences of loss of function in *CIAO1*, the gene encoding a key component of the CIA complex, have thus far been unknown as its role in the biogenesis of Fe-S clusters for nucleocytoplasmic Fe-S enzymes has been inferred based solely on knockdown experiments in cultured cells. ^17, 31^ We report here that biallelic pathogenic variants in *CIAO1* cause a novel disorder in human with predominantly neuromuscular but also multisystemic manifestations. The amino acid sequence of CIAO1 is highly conserved across species from human to zebrafish as are the residues mutated in the patients reported here (Fig. S2). The complementary studies in patient-derived cell lines and the biochemical characterization of the *CIAO1* variants confirmed the deleterious nature of the variants, which caused protein instability and compromised interaction with other CIA components and with Fe-S recipient proteins. Interestingly, in cell lines derived from patients, the chronic depletion of CIAO1 was associated with a concomitant decrease in the levels of its established interacting partners, FAM96B, MMS19, and FAM96A. This outcome is unprecedented, as previous reports on short-interfering RNA (si-RNA) mediated knockdown (KD) of *CIAO1* did not indicate a loss of stability in FAM96B, MMS19, or FAM96A^31, 34^. Our findings highlight the distinction between the impact of an acute, temporary depletion of CIAO1, as achieved by si-RNA-mediated KD, and a sustained loss, as observed in patient-derived cell lines, emphasizing the predominant involvement of the CIA components in a shared cellular pathway. Suboptimal levels of the CIA machinery led to compromised biogenesis of multiple Fe-S enzymes that play critical roles in genome maintenance (such as RTEL1), DNA replication (POLD1), tRNA modifications (ELP3 and CDKAL1), and purine and pyrimidine metabolism (PPAT and DPYD, respectively).

Patients presented with slowly progressing proximal and axial muscle weakness, respiratory insufficiency, elevated CK, and histologic features consistent with a dystrophic myopathy characterized by abnormal mitochondrial morphology. Additional clinical manifestations of note included learning disabilities and neurobehavioral comorbidities, iron deposition in brain deep nuclei, macrocytic anemia, and gastrointestinal symptoms, indicating multisystemic consequences of diminished CIAO1 activity. The age of recognition of myopathic symptoms varied among our patients from early childhood to adolescence. Interestingly, this variability was also observed among the two patients carrying the same biallelic variants in *CIAO1* (P2 and P3). Three of four patients (P1, P2, P4) remain ambulant, while P3 has progressed to become wheelchair bound in her 20s, implying that the rate of progression of muscle weakness may vary as well. In this study, carriers, including the father of P1, were not clinically affected, indicating that one functional copy of *CIAO1* is sufficient for normal physiology which is also consistent with the variants being only in heterozygous state in the GnomAD database. The *CIAO1* variants identified in our patients result in only a partial loss of function, as global knockout is likely to be incompatible with life. Thus far, only genetic ablation of MMS19 has been attempted in mice, leading to preimplantation death. ^15^

Interestingly, the pattern of brain iron deposition observed in the *CIAO1*-deficient patients markedly resembles that of patients with Neurodegeneration with Brain Iron Accumulation (NBIA), a clinically and genetically heterogenous group of disorders affecting children and adults. ^35^ NBIA is often first suspected when increased basal ganglia iron is observed on brain magnetic resonance imaging. Multisystem involvement has also been described for NBIA patients and may extend to hair loss, diabetes, hearing loss, gonadal dysfunction, and intellectual disabilities, while muscle involvement has not been explicitly reported as part of the disorder. ^35^ The broad phenotypic manifestation of loss of function in *CIAO1* underscores the critical role of the CIA machinery in delivering Fe-S cofactors to numerous essential nuclear and cytoplasmic Fe-S enzymes involved in all aspects of DNA metabolism, tRNA modification and protein translation. Given the ubiquitous nature of these processes, it is likely that the spectrum of *CIAO1*-related disorders may vary and expand as new pathogenic variants are ascertained, depending on the specific effect that the amino acid substitutions have on protein stability and function.

Loss of function in several components of the Fe-S biogenesis pathway has been linked to multiple rare human conditions that manifest with different patterns of systemic or tissue-specific involvement.^27^ Relevant examples are the multiple mitochondrial dysfunctions syndromes (OMIM #605711, #614299, #615330, #616370, #617613, #617954, #620423), which manifest as severe autosomal recessive disorders of systemic energy metabolism, resulting in weakness, respiratory failure, severely impaired neurologic development, lactic acidosis, and early death. ^27^ However, several studies have also identified distinctive tissue-specific manifestations as the main characteristic in a number of disorders caused by mutations in the Fe-S biogenesis components, including the sideroblastic anemia caused by loss of function in *GLRX5* (OMIM # 616860).^36, 37^ Additionally, a subset of muscle specific disorders has also been documented, including the myopathy with lactic acidosis due to aberrant splicing of the *ISCU* transcript (OMIM # 255125).^38^ This condition leads to a muscle specific loss of Fe-S proteins, along with mitochondrial iron accumulation, causing symptoms such as poor endurance, muscle cramps, lactic acidosis and severe episodes of myoglobinuria. ^38, 39^ While global knock-out of *ISCU* in mice results in early embryonic lethality, ^40^ the intronic variant identified in patients allows an aberrant splicing pattern of *ISCU* that leaves some residual function, potentially providing valuable insights into the muscle specific phenotype of the disease. ^38, 41^

*FDX2*, encoding a small protein involved, along with FDXR, in donating electrons to nascent Fe-S clusters, was the second gene in the iron-sulfur biogenesis pathway to be linked to a mitochondrial myopathy (OMIM #251900).^42^ Following the first report in 2014 of a single patient who presented with adolescent onset of autosomal recessive mitochondrial myopathy,^42^ a second report of a novel variant, five years later, broadened the spectrum of the *FDX2*-related disorder in six patients from two Brazilian families.^43^ This new variant was linked to early onset of neurological symptoms, optic atrophy and myopathy characterized by recurrent episodes of cramps, myalgia and muscle weakness.^43^ Sensory-motor axonal neuropathy and leukoencephalopathy with reversible white matter changes were also shown to be part of the extended phenotype.^43^

The muscle histopathological and ultrastructural features of the *CIAO1*-deficient patients demonstrate a combination of unique characteristics, including mixed moderate myopathic and dystrophic changes. Additionally, strikingly large and morphologically abnormal mitochondria were observed, while histopathology lacked classic findings commonly seen in mitochondrial myopathies such as COX-negative fibers and SDH deficiency. This distinction sets the CIAO1 myopathy apart from the classical mitochondrial Fe-S-associated myopathies like the ISCU– and FDX2-myopathies. While the CIAO1-related muscle pathology does not fit the conventional criteria of a mitochondrial myopathy, it exhibits a discernable mitochondrial dysfunction, as demonstrated by histological, ultra-structural, and functional assessments. We speculate that the mitochondrial dysfunction in CIAO1-deficient muscle might be secondary to the loss of several nucleocytoplasmic Fe-S enzymes that depend on CIA for function. Sufficient levels of those enzymes are critical to meet the cellular needs for transcriptional and translational activities. Therefore, this impairment becomes particularly notable in muscle tissue, known for its high protein turnover rates. Myofibers contain in fact hundreds to thousands of myonuclei which are responsible for ongoing transcription of muscle-specific genes coding for contractile elements, extracellular matrix components, and subunits of the mitochondrial oxidative phosphorylation system.^44^ The defective mitochondrial function in muscle, which is not detectable in fibroblasts, supports the notion of an increased demand for transcriptional and translational activities in muscle tissue, which is contingent upon maintaining adequate levels of nucleocytoplasmic Fe-S enzymes. Interestingly, the enlarged mitochondria were observed mainly in type 1 myofibers (Figure 2F, type 1 = darker myofibers), which are known to be rich in mitochondria and to rely on aerobic metabolism, further supporting the idea of a cell specific threshold requirement of the CIA machinery for proper function.

Although the exact phenotypic spectrum of the *CIAO1*-related disorder remains to be fully defined and will likely become clearer as more patients are identified, our findings contribute to a better understanding of the role of CIAO1 in the biogenesis of Fe-S clusters for nucleocytoplasmic Fe-S enzymes. Additionally, our study defines the essential role of CIAO1 in vivo for human health and offers insights into a novel multisystemic disorder.

## Abbreviations

CIA: Cytoplasmic Iron-sulfur Assembly
CK: Creatine Kinase
CNS: central nervous system
COX: Cytochrome c Oxidase
EM: Electron Microscopy
FVC: forced vital capacity
H&E: Hematoxylin and Eosin
ISC(s): Iron Sulfur Cluster(s)
NADH-TR: Nicotinamide Adenine Dinucleotide Tetrazolium Reductase
QSM: Quantitative Susceptibility Mapping
SDH: Succinic Dehydrogenase
SWI: Susceptibility Weighted Imaging

## Acknowledgements

The authors want to thank the patients and their families for their willingness to contribute, and Christopher Mendoza, Kia Brooks, Angela Kokkinis, Dr. Abdullah Alqahtani and Gilberto (“Mike”) Averion for their help in clinical evaluations and samples collection.

We thank Darren Chambers and Andrew Dawson for their help with the EM images.

The authors thank the NIHR Biomedical Research Centre at Great Ormond Street for support to the Biobank at the Dubowitz Neuromuscular Centre and the NICHD Intramural Program for support.

Figures 1B and 2A created with BioRender.com.

