## Supplemental Information for "Loss of Function of the Cytoplasmic Fe-S Assembly Protein CIAO1 Causes a Neuromuscular Disorder with Compromise of Nucleocytoplasmic Fe-S Enzymes"

### Inventory of Supplemental Information

#### Supplemental figures

**Figure S1**. RNA sequencing, P1 fibroblasts

**Figure S2**. Multiple sequence alignment of CIAO1 amino acid sequences and location of the amino acid residues altered in patients.

**Figure S3**. The CIAO1 variants identified in patients have greatly diminished stability compared to wild type CIAO1 and impaired binding to the components of the Fe-S biogenesis machinery and recipient apo-proteins.

**Figure S4**. Iron homeostasis is maintained in patient-derived cells because of two opposing regulatory axes that are at equilibrium.

**Figure S5**. P1-derived fibroblasts do not exhibit a profound mitochondrial defect.

**Figure S6**. Levels of the regulators of mitochondrial dynamics, OPA1 and mitofusin 1 and 2 (MFN1/2), were unaltered in P1- derived fibroblasts.

#### Supplemental Tables

**Table S1. Predicted pathogenicity of *CIAO1* variants identified in patients.**

#### Supplemental references

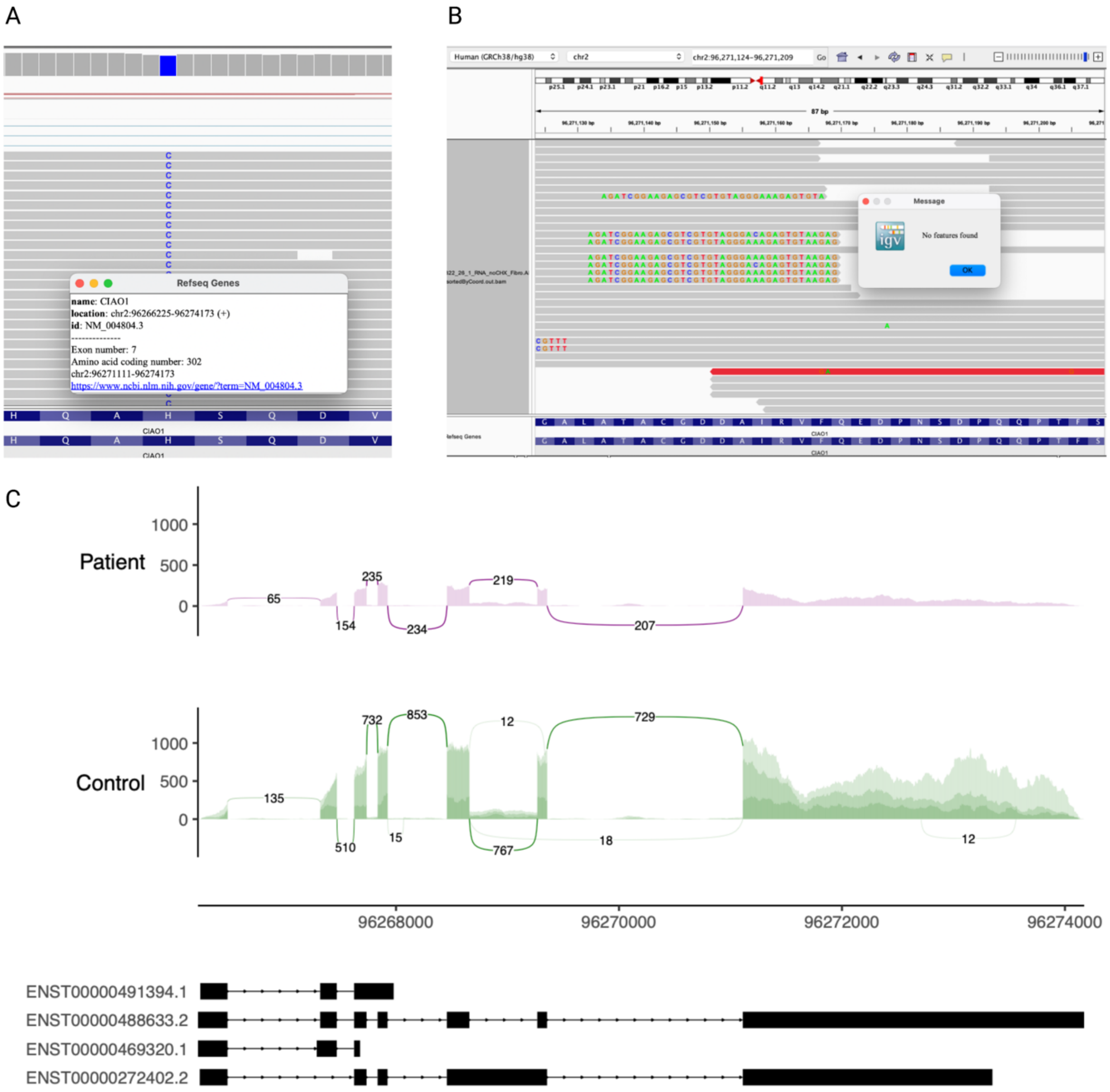


**Figure S1. RNA sequencing, P1 fibroblasts. A.** Maternal allele is homozygous in P1 RNAseq sample **B.** Soft clipping in exon7 do not map, showing there is no evidence for paternal reads **C.** Sashimi plots comparing fibroblasts CIAO1 sequencing reads in P1 (purple) and three control fibroblasts samples (green)

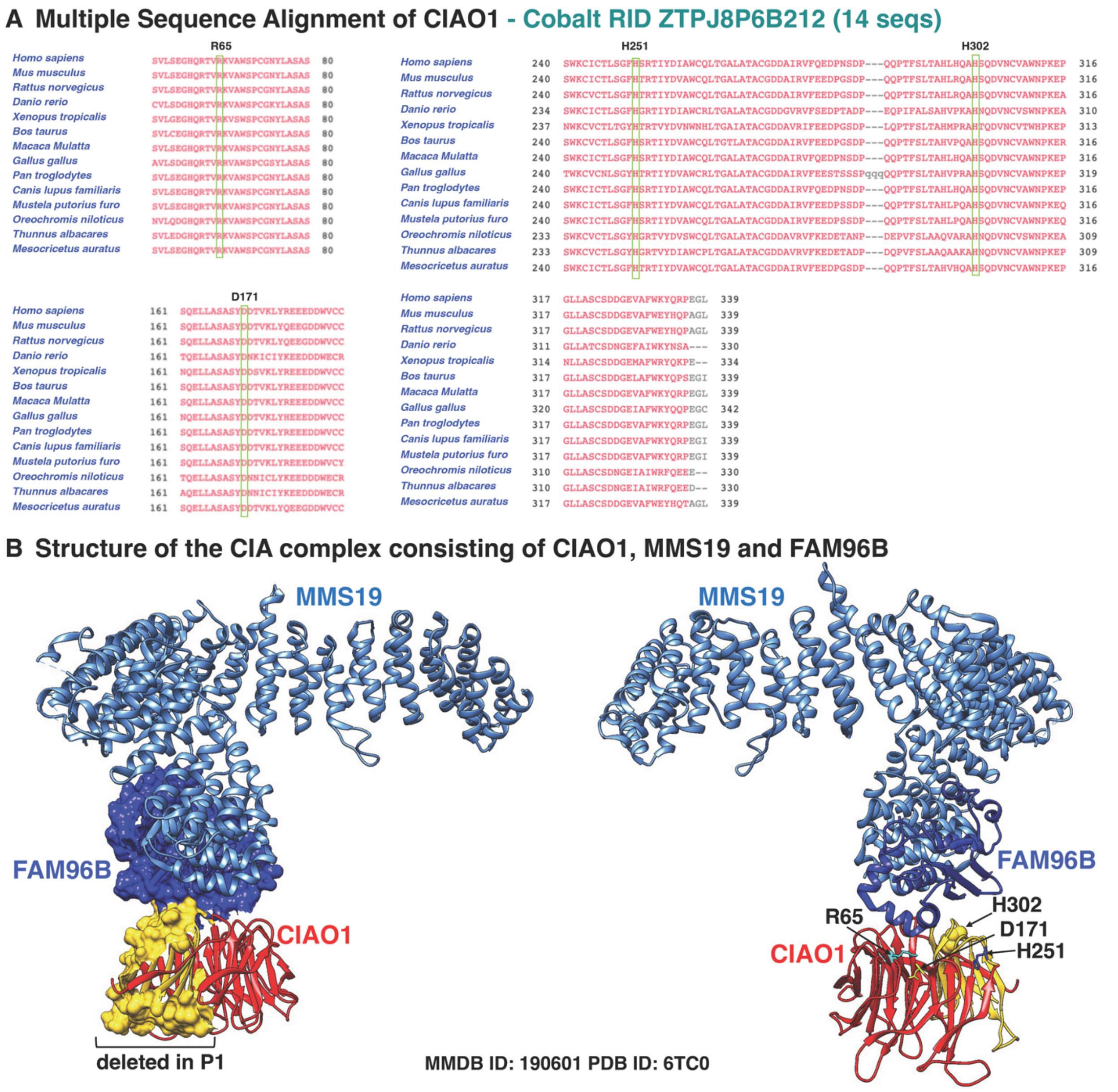


**Figure S2. Multiple sequence alignment of CIAO1 amino acid sequences and location of the amino acid residues altered in patients. A.** Multiple sequence alignment of CIAO1 sequences generated with Constraint-based Multiple Alignment Tool (COBALT, NCBI) showing complete conservation across different species of the amino acid residues altered in the patients. **B.** 3D structure of the CIA complex consisting of CIAO1, MMS19 and FAM96B (PDB ID: 6TC0). On the left, CIAO1 is shown in ribbon- and surface- mode representations and colored in red, except for the domain deleted in P1 which is rendered in surface-mode representation and colored in yellow. On the right, CIAO1 is shown in ribbon-mode representation and the location of amino acid residues altered in the patients are labeled and pointed by arrows.

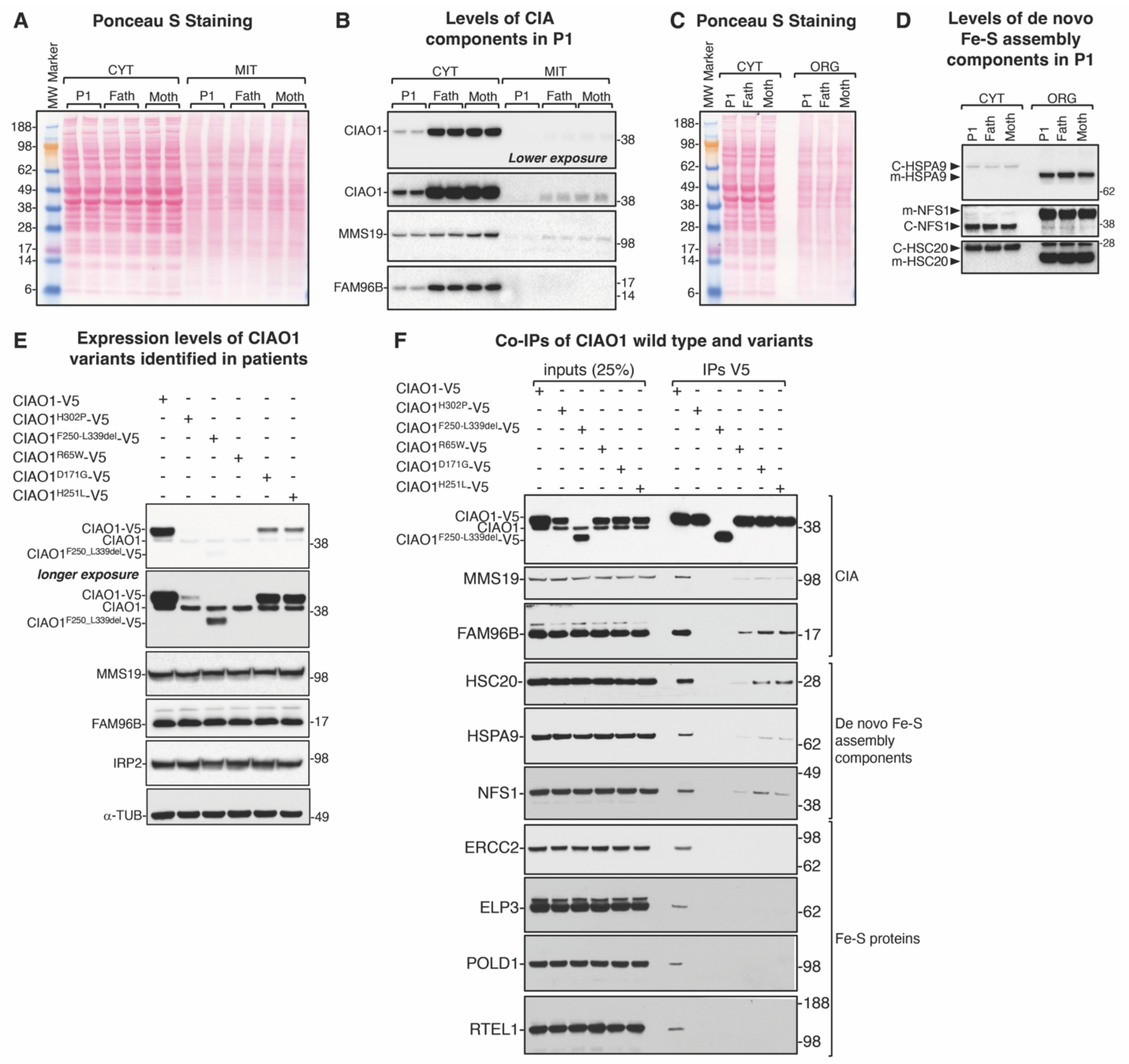


**Figure S3. The *CIAO1* variants identified in patients have greatly diminished stability compared to wild type CIAO1 and impaired binding to the components of the Fe-S biogenesis machinery and recipient apo-proteins. A.** Ponceau S staining of nitrocellulose membrane allows visualization of proteins extracted from cytosolic (CYT) and mitochondrial (MIT) lysates obtained from P1- and parental- derived fibroblasts. MW marker (molecular weight marker). **B.** Immunoblots to components of the CIA machinery shows specific cytosolic (but not mitochondrial) localization of CIAO1, MMS19 and FAM96B. Levels of these components were profoundly diminished in P1 cytosolic lysates compared to parental derived fibroblasts. **C.** Ponceau S staining of nitrocellulose membrane allows visualization of proteins extracted from cytosolic (CYT) and mitochondrial (MIT) lysates obtained from P1- and parental-derived fibroblasts. **D.** Immunoblots to components of the *de novo* Fe-S cluster biogenesis machinery shows dual localization to cytosol (CYT) and mitochondria (MIT) of HSPA9, HSC20 and NFS1, consistent with previously reported results [1-3]. **E.** Immunoblots to components of the CIA pathway, CIAO1, MMS19 and FAM96B, Iron Regulatory Protein 2 (IRP2) and loading control ($\alpha$-TUB) in HeLa cells transiently transfected with C-terminally V5-tagged *CIAO1* wild type and variants identified in patients, as indicated. All *CIAO1* variants exhibited profoundly diminished stability 16 hours post-transfection. **F.** Co-immunoprecipitation (co-IP) experiments of recombinantly expressed V5-tagged *CIAO1* wild type and variants identified in patients, as indicated. In order to normalize for the reduced stability of the CIAO1-V5 variants (presented in panel E), the V5 agarose beads incubated with the lysates obtained from cells expressing the variants were recovered in 25µl of elution buffer (EB), whereas 65µl of EB were used for the wild type sample. Additionally, five times more IP eluate was loaded onto the gel for the variants. Lysates were prepared 8 hours post-transfection.

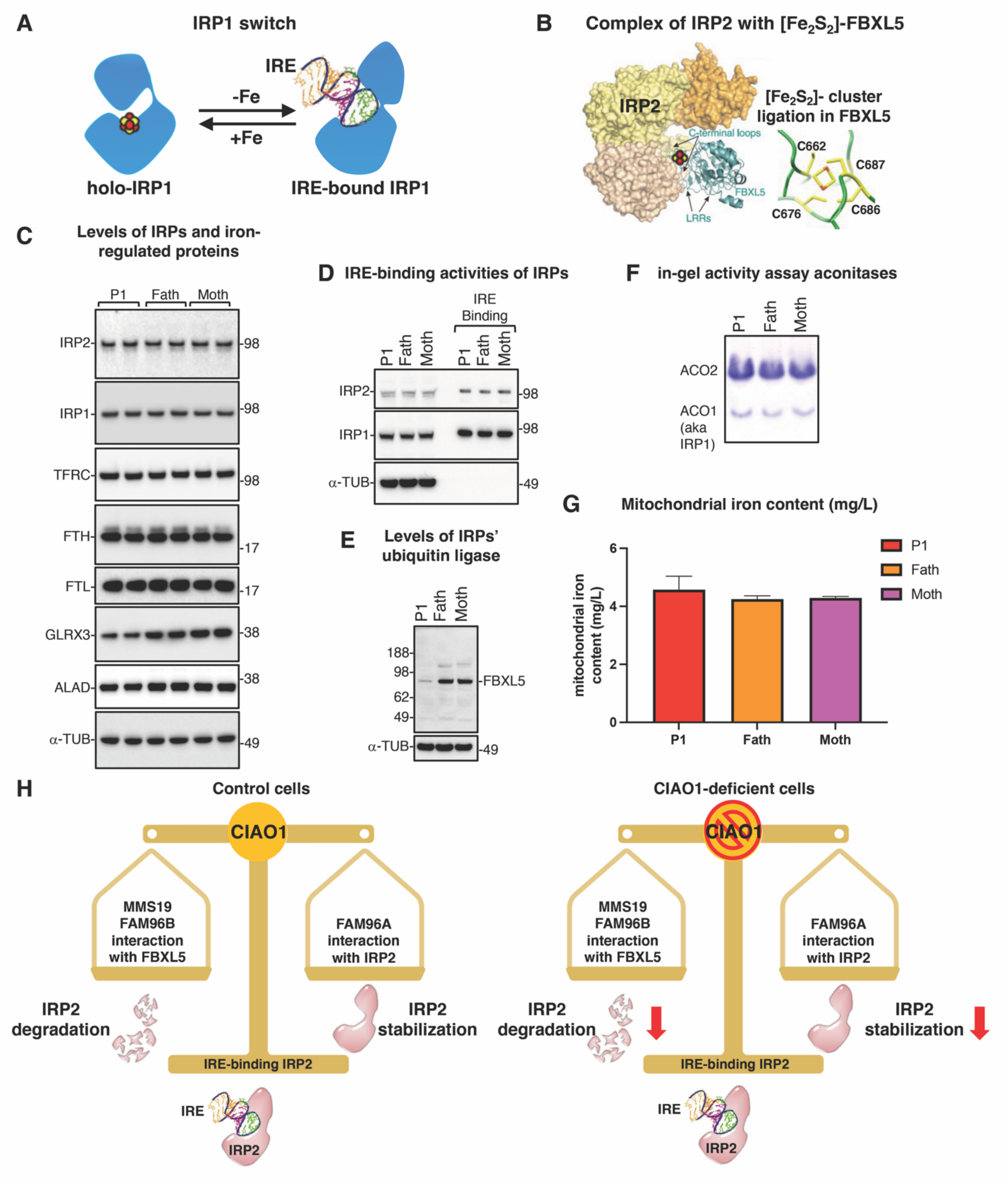


**Figure S4. Iron homeostasis is maintained in patient-derived cells because of two opposing regulatory axes that are at equilibrium. A.** IRP1, a protein with dual function. IRP1 alternates between a cytosolic aconitase holo-form when it ligates a $[Fe_4S_4]$ cluster in its active site cleft under iron replete conditions (+Fe) and an apo-protein that lacks the cluster and binds to Iron Responsive Element (IRE) stem-loop structures present in several transcripts encoding iron metabolism proteins under iron deficiency (-Fe). Upon binding, IRP1 represses translation of transcripts that contain IREs near the 5'-end (e.g., ferritin H and L) and stabilizes from endonucleolytic degradation mRNAs that contain IREs at the 3'-UTR (e.g., transferrin receptor). **B.** Complex of IRP2 with its ubiquitin-ligate FBXL5. FBXL5 has been reported to interact with the CIA complex[4] and to ligate a [$Fe_2S_2$] cluster[5]. **C.** Immunoblots on lysates from P1- and parental- derived fibroblasts to Iron Regulatory Proteins 1 and 2 (IRP1/2), transferrin receptor (TFRC), ferritin H and L (FTH and FTL), glutaredoxin 3, and the heme biosynthetic enzyme ALAD. Alpha-tubulin ($\alpha$-TUB) was used as a reference for loading control. **D.** IRE-binding activities of IRP1 and IRP2 in P1- and parental-derived fibroblasts. **E.** Levels of the E3-ubiquitin ligase, FBXL5, that degrades IRPs under iron replete conditions. **F.** In-gel activity assays of cytosolic (ACO1) and mitochondrial (ACO2) aconitases demonstrated unaltered activities of the [$Fe_4S_4$] enzymes in the patient-derived cells (P1) compared to control fibroblasts (parental cells). **G.** Iron content in P1- and parental-derived mitochondria as assessed by inductively coupled plasma mass spectrometry (ICP-MS) ($n$=3 biological replicates). **H.** Model depicting the two opposing regulatory axes which control maintenance of IRP2 protein levels and iron homeostasis in the CIAO1-deficient patient-derived cells. Two multi-protein complexes that share CIAO1 as a component control IRP2 protein levels and cellular iron homeostasis. In control cells (on the left), levels of IRP2 result from a balance of two opposing regulatory axes: degradation of IRP2 by FBXL5 and stabilization of IRP2 through its interaction with the CIAO1/FAM96A complex. Under steady state conditions:

1. IRP2 is degraded by FBXL5 whose levels and ubiquitin-ligase activity depend on its ability to interact with CIAO1, FAM96B and MMS19[4] (left arm of the balance);

2. IRP2 is stabilized by its interaction with the CIAO1/FAM96A complex[6] (right arm of the balance).

In the patient-derived cells (on the right), loss of CIAO1 prevents the regular turnover of IRP2 (left arm of the balance) because of the decreased levels of FBXL5. Loss of FBXL5 would be expected to cause an increase in IRP2 protein levels. However, because of the compromised stabilization of IRP2 due to loss of CIAO1 and FAM96A, levels of IRP2 remain unchanged in the patient-derived cells compared to control. The rate of IRP2 degradation is decreased in the patient-derived cells, while its stabilization is concomitantly impaired, resulting in no significant change in IRP2 levels under steady state conditions.

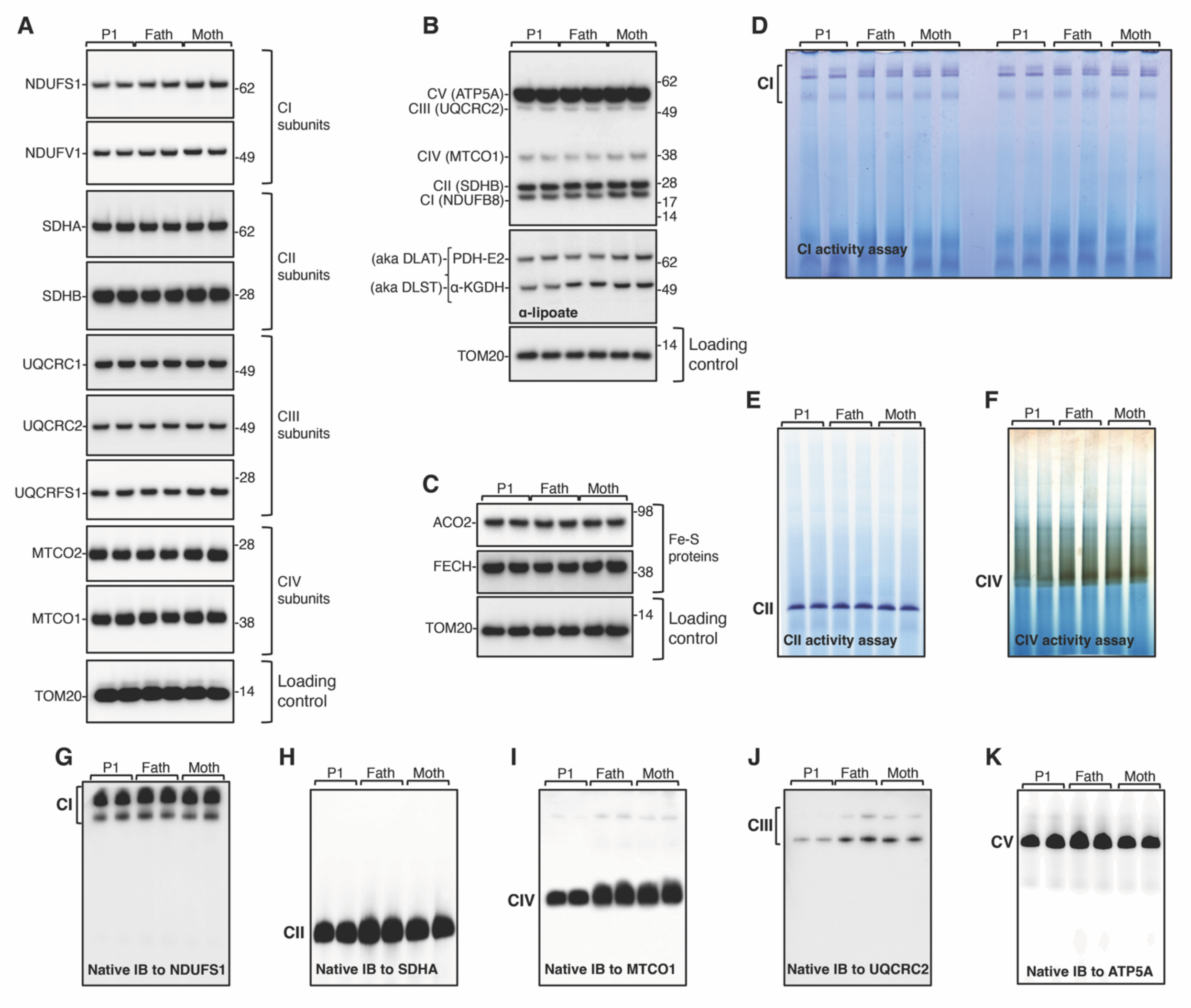


**Figure S5. P1-derived fibroblasts do not exhibit a profound mitochondrial defect. A.** SDS immunoblots to subunits of mitochondrial respiratory complex I (NDUFS1, NDUFV1), complex II (SDHA, SDHB), complex III (UQCRC1, UQCRC2, UQCRFS1), and complex IV (MTCO1, MTCO2) in lysates obtained from P1- and parental- derived fibroblasts. Levels of TOM20 are shown as a reference for loading control. **B.** SDS immunoblots to total oxidative phosphorylation subunits, as indicated, and to lipoate in P1- and parental- derived cells. **C.** SDS immunoblots to the mitochondrial Fe-S cluster subunits aconitase (ACO2) and to the terminal heme biosynthetic enzyme ferrochelatase (FECH) in P1- and parental- derived fibroblasts. **D.** In-gel NADH oxidase (diaphorase) activity assay of mitochondrial complex I (CI) in P1- and parental-derived cells. **E.** In-gel succinate dehydrogenase (CII) activity assay in P1- and parental- derived cells. **F.** In-gel cytochrome c oxidase (CIV) activity assay in P1- and parental- derived fibroblasts. **G-K.** Native immunoblots to NDUFS1 (complex I subunit), SDHA (complex II subunit), MTCO1 (complex IV subunit), UQCRC2 (complex III subunit) and ATP5A (complex V subunit), respectively, in P1- and parental-derived fibroblasts to assess the overall levels of fully assembled respiratory complexes.

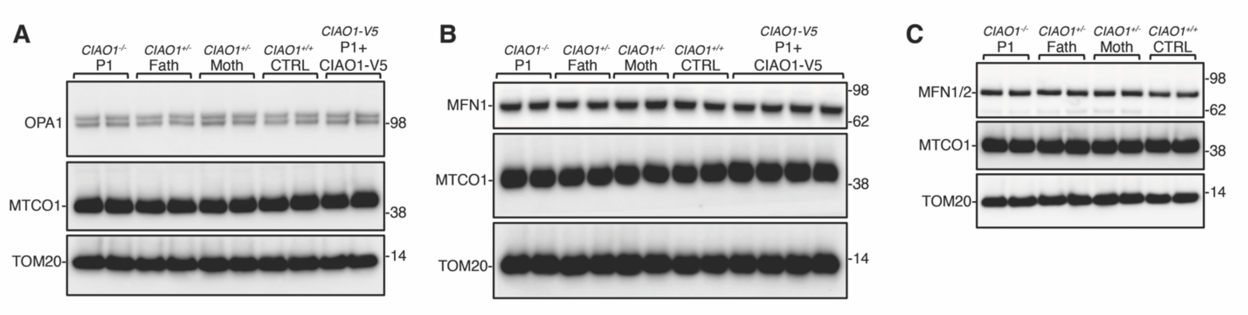


**Figure S6. Levels of the regulators of mitochondrial dynamics, OPA1 and mitofusin 1 and 2 (MFN1/2), were unaltered in P1- derived fibroblasts. A.** Immunoblots to OPA1 and the complex IV subunit MTCO1 in P1-, parental-, control- (corresponding to fibroblasts expressing two wild type copies of *CIAO1*) fibroblasts and in P1- derived fibroblasts that had been lentivirally transduced with V5-tagged *CIAO1* wild type. **B.** Immunoblots to MFN1 and the complex IV subunit MTCO1 in P1-, parental-, control- (corresponding to fibroblasts expressing two wild type copies of *CIAO1*) fibroblasts and in P1- derived fibroblasts that had been lentivirally transduced with V5-tagged *CIAO1* wild type. **C.** Immunoblots to MFN1, MFN2 and the complex IV subunit MTCO1 in P1-, parental-, control- (corresponding to fibroblasts expressing two wild type copies of *CIAO1*) fibroblasts and in P1- derived fibroblasts that had been lentivirally transduced with V5-tagged *CIAO1* wild type. In **A-C**, levels of TOM20 are presented as a reference for loading control.

#### Table S1. Predicted pathogenicity of *CIAO1* variants identified in patients.

| Chromosomal location[1] | *CIAO1* Variant[2] | Patients identifiers (number of alleles) | gnomAD[3] | gnomAD – number of homozygous[3] | ExAC2[4] | REVEL[5] | Polyphen 2[6] | SIFT[7] |
|---|---|---|---|---|---|---|---|---|
| chr2-96936974 | c.905A>C p.His302Pro | P1, P2, P3 (n = 3) | 0.0000212 1 | 0 | 0.0000082 46 | Deleterious (Strong) (0.95) | Deleterious (Moderate) (1) | Uncertain (0.003) |
| chr2-96933112 | c.193C>T p.Arg65Trp | P2, P3 (n = 2) | 0.001011 | 0 | 0.0009243 | Uncertain (0.45) | Deleterious (Moderate) (1) | Deleterious (Supporting) (0) |
| chr2-96934217 | c.512A>G p.Asp171Gly | P4 (n = 1) | 0 | n/a | 0 | Deleterious (Moderate) (0.93) | n/a | Deleterious (Supporting) (0) |
| chr2-96935066 | c.752A>T p.His251Leu | P4 (n = 1) | 0.0000039 76 | 0 | 0.0000082 37 | Deleterious (Moderate) (0.92) | Deleterious (Supporting) (1) | Uncertain (0.002) |
| chr2:96936630-96937369 | DEL: chr2:96936630-96937369 p.Phe250_Leu339del | P1 (n=1) | 0 | n/a | 0 | n/a | n/a | n/a |

[1]reference sequence: NM_004804.3
[2]hg19
[3]gnomAD v2.1.1: 141,456 samples
[4]ExAC v1.0: 60,706 samples
[5]REVEL, Ensemble Method for Predicting the Pathogenicity of Rare Missense Variant based on a combination of scores from 13 individual tools; https://sites.google.com/site/revelgenomics/
[6]Polyphen2, predicts possible impact of an amino acid substitution on the structure and function of a human protein; http://genetics.bwh.harvard.edu/pph2/
[7]SIFT, predicts whether an amino acid substitution affects protein function based on sequence homology and the physical properties of amino acids; https://sift.bii.a-star.edu.sg/
